# Target trial emulation for a two-arm, parallel, stratified pragmatic RCT of CABG versus PCI in people with high-risk characteristics requiring myocardial revascularisation (High-Risk REVASC)

**DOI:** 10.64898/2026.04.30.26352120

**Authors:** Weiqi Liao, Ian R White, Rebecca M Turner, Wenyue Li, Muhammad Rashid, Cassandra L Brookes, Shaun Barber, Guillaume Marquis-Gravel, Mark C Petrie, National Cardiac Surgery Patient and Public Involvement Group, Jean Rouleau, Stephen Fremes, Gavin J Murphy

## Abstract

**Aim:** This study aimed to investigate whether treatment effects (TE) for coronary artery bypass grafting (CABG) versus percutaneous coronary intervention (PCI) were consistent for people requiring coronary revascularisation with high-risk characteristics.

**Methods:** We used target trial emulation (TTE) as study design and observational data from Hospital Episode Statistics (HES). The target population was people requiring multivessel coronary revascularisation with at least one of seven high-risk characteristics: age >75 years, female, heart failure, chronic kidney disease, peripheral vascular disease, intermediate frailty risk, or presentation with acute coronary syndrome. The intervention was CABG and the comparison was PCI. Outcomes included all-cause and cardiovascular (CV) mortality, CV hospitalisation, and major adverse cardiovascular events within 5 years of the index procedure. This study included four research stages: (1) latent class analysis (LCA) to identify mutually exclusive patient clusters representing different clinical phenotypes, (2) instrumental variable analysis (IVA) to estimate the TE for the whole population and each patient class; (3) repeating IVA in an emulated trial population obtained by matching a previous cardiac surgery trial enriched for high-risk characteristics to the HES population; (4) co-designed a pragmatic randomised controlled trials (RCT) with multiple stakeholders to address uncertainty identified from the analyses above.

**Results:** Of 103,670 patients in the target population, 62,048 (59.9%) received CABG and 41,622 (40.1%) received PCI. Seven patient classes were identified as the best solution from LCA. The emulated trial consisted of 3124 patients, 1,588 (50.8%) in CABG arm and 1,536 (49.2%) in PCI arm. Patients aged >75 years without CKD (Class 1) who received CABG showed consistent benefits in all primary and secondary outcomes. For the other six patient classes, heterogeneity in benefits and harms between CABG and PCI existed both between and within classes in clinical outcomes. An RCT to resolve the remaining uncertainty using Bayesian approach would require 3000 participants to detect a hazard ratio of 0.7 with a family-wise type 1 error rate <5% and 90% power across all seven classes.

**Conclusion:** TTE of coronary revascularisation options in people with high-risk characteristics demonstrated mixed benefits and harms both between and within disease phenotypes. A trial to address uncertainty as to the indications for CABG versus PCI in this target population is feasible.

**Study registration:** ClinicalTrials.gov (NCT05853536, https://clinicaltrials.gov/study/NCT05853536)

## Introduction

Treatment guidelines for myocardial revascularisation in multivessel coronary artery disease (CAD) recommend coronary artery bypass grafting (CABG) or percutaneous coronary intervention (PCI) based on the severity and extent of coronary atheroma, and the presence or absence of comorbidities such as diabetes and left ventricular dysfunction.^1,2^ Guidelines make no specific references to common high-risk characteristics including older age (>75 years), female sex, comorbidities such as chronic kidney disease (CKD), peripheral vascular disease (PVD), and frailty, or presentation with acute coronary syndrome (ACS). These risk factors are considered in treatment decisions as important determinants of clinical outcomes^3^, but were underrepresented or even excluded from existing randomised controlled trials (RCTs) of CABG versus PCI.^4,5^ Managing these patients in the absence of evidence leads to unwarranted variation in treatment choices and outcomes. ^6^ The aim of this study was to address this uncertainty using target trial emulation (TTE).

An RCT of CABG versus PCI in high-risk groups presents specific design challenges. First, high-risk groups demonstrate significant overlap or interdependence.^7^ Trials restricted to individual risk factors do not address this clinical complexity; multiple two-arm parallel group trials that consider each high-risk characteristic individually would take many years to complete. Due to the degree of overlap of these conditions within individuals, trial analyses demonstrating futility or effectiveness for one high-risk characteristic would affect equipoise in other groups where they are overrepresented. Second, people with different high-risk characteristics demonstrate different disease trajectories, as well as different interactions with the mode of revascularisation. ^8,9^ Third, as the number of high-risk characteristics increases, the size of the treatment effect of the more invasive CABG intervention required to change practice may also increase.

We addressed this complexity in four research stages. First, to address heterogeneity and interdependence of high-risk characteristics, we used latent class analysis (LCA, an unsupervised machine learning method) to identify non-overlapping patient clusters for stratification. Second, we used instrumental variable analysis (IVA, causal inference) to estimate the treatment effects (TE) for different clinical outcomes between the two modes of revascularisation (CABG versus PCI) for the whole study population and each patient cluster. Third, to emulate TEs in a likely clinical trial population, we repeated IVA in a population matched to individual participant data (IPD) from a clinical trial in people with high-risk characteristics undergoing surgical revascularisation recruited in UK centres. Finally, we used these results to co-design an efficient, pragmatic RCT of CABG versus PCI in high-risk populations in partnership with patients, the public, and stakeholders that will address remaining uncertainty.

## Methods

This study was approved by the University of Leicester Research Ethics Committee and NHS Digital (Ref: 22322-yll15-ls:cardiovascularsciences) and was prospectively registered (NCT05853536). We followed the TARGET statement^10^ (Transparent Reporting of Observational Studies Emulating a Target Trial) to design, conduct, and report this study (schecklist in the supplementary materials). Data management and statistical analyses were conducted in Stata 19 and R 4.5.1.

The study protocol, including a detailed statistical analysis plan, has been published. ^11^ We used the Hospital Episodes Statistics Admitted Patient Care (HES-APC, 2007-2020) as the main data source. The **<u>target population</u>** was adult patients with at least one high-risk characteristic admitted to National Health Service (NHS) hospitals in England from 1 April 2009 to 31 March 2015 who underwent isolated CABG or multivessel PCI, with a complete 5-year follow-up to 31 March 2020, representing routine healthcare before COVID-19. The **<u>intervention</u>** procedure was CABG and the **<u>comparator</u>** was multivessel PCI. Diagnoses and procedures within 2 years prior to the index procedure were used to establish patients’ medical histories and comorbidities, where the index procedure meant the date of each patient received CABG or multivessel PCI (“**<u>Time zero</u>**”). The composite primary **<u>outcome</u>** was all-cause death or cardiovascular rehospitalisation within 5 years of the index revascularisation procedure. Secondary outcomes included all-cause death, cardiovascular death, cardiovascular rehospitalisation due to major adverse cardiovascular events (MACE); myocardial infarction (MI), heart failure (HF) and stroke within 5 years of the index procedure. Detailed information on PICO and all study design components for both target and emulated trials is in **Table 1**. Participant flow diagram is in **Figure 1**. The conceptual framework of the study is in **Figure S1**.

**Table 1.**
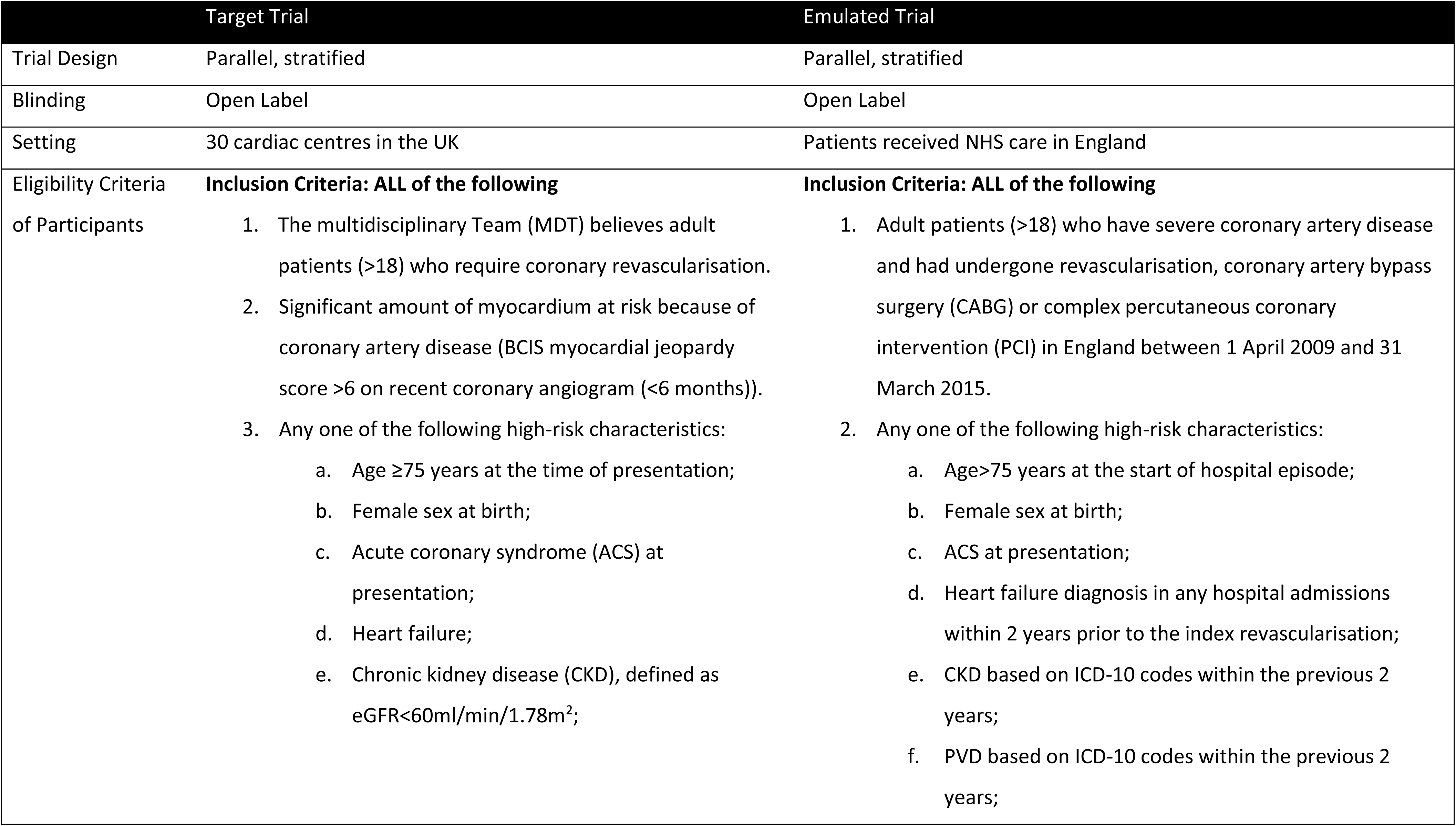
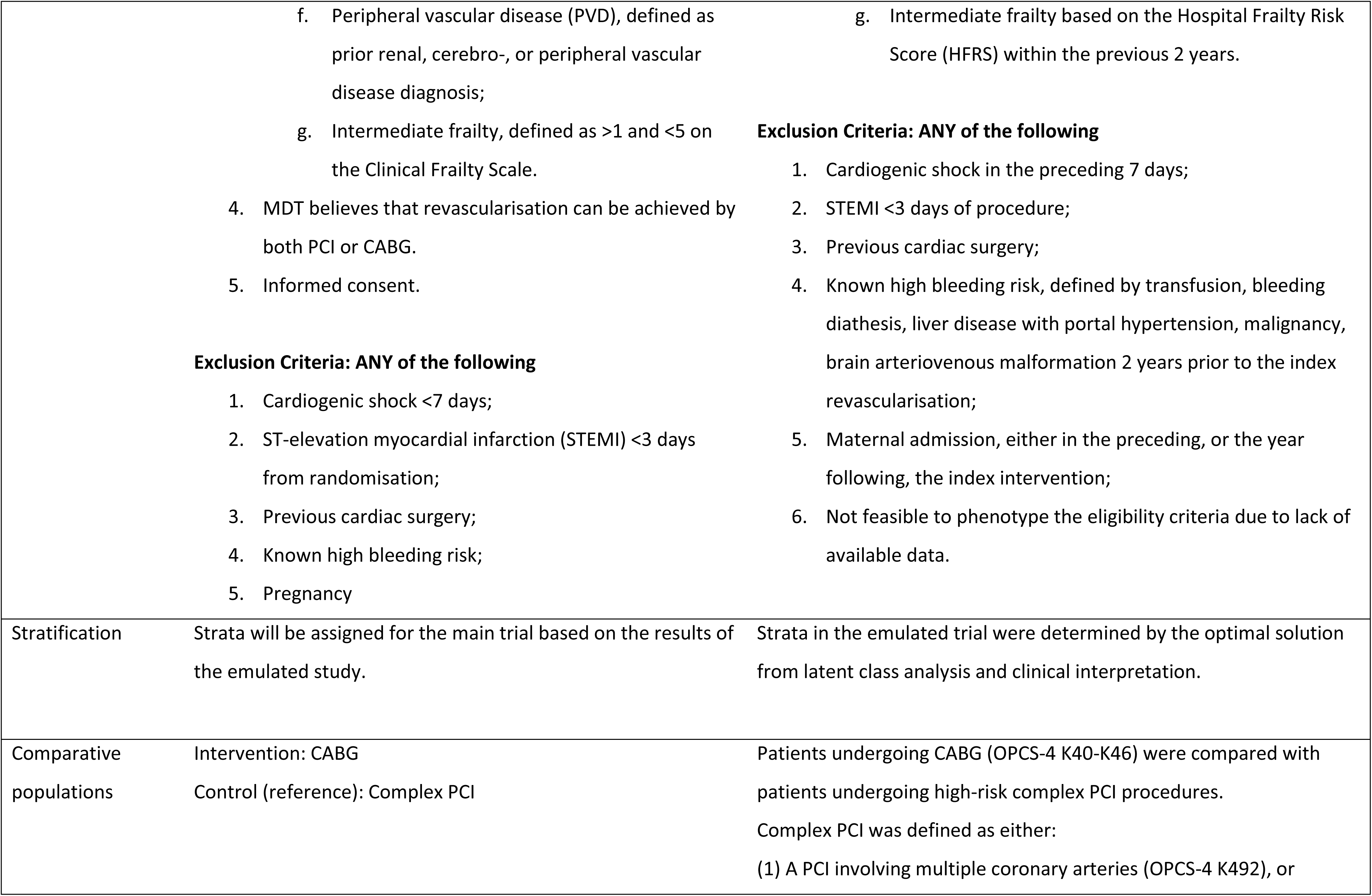
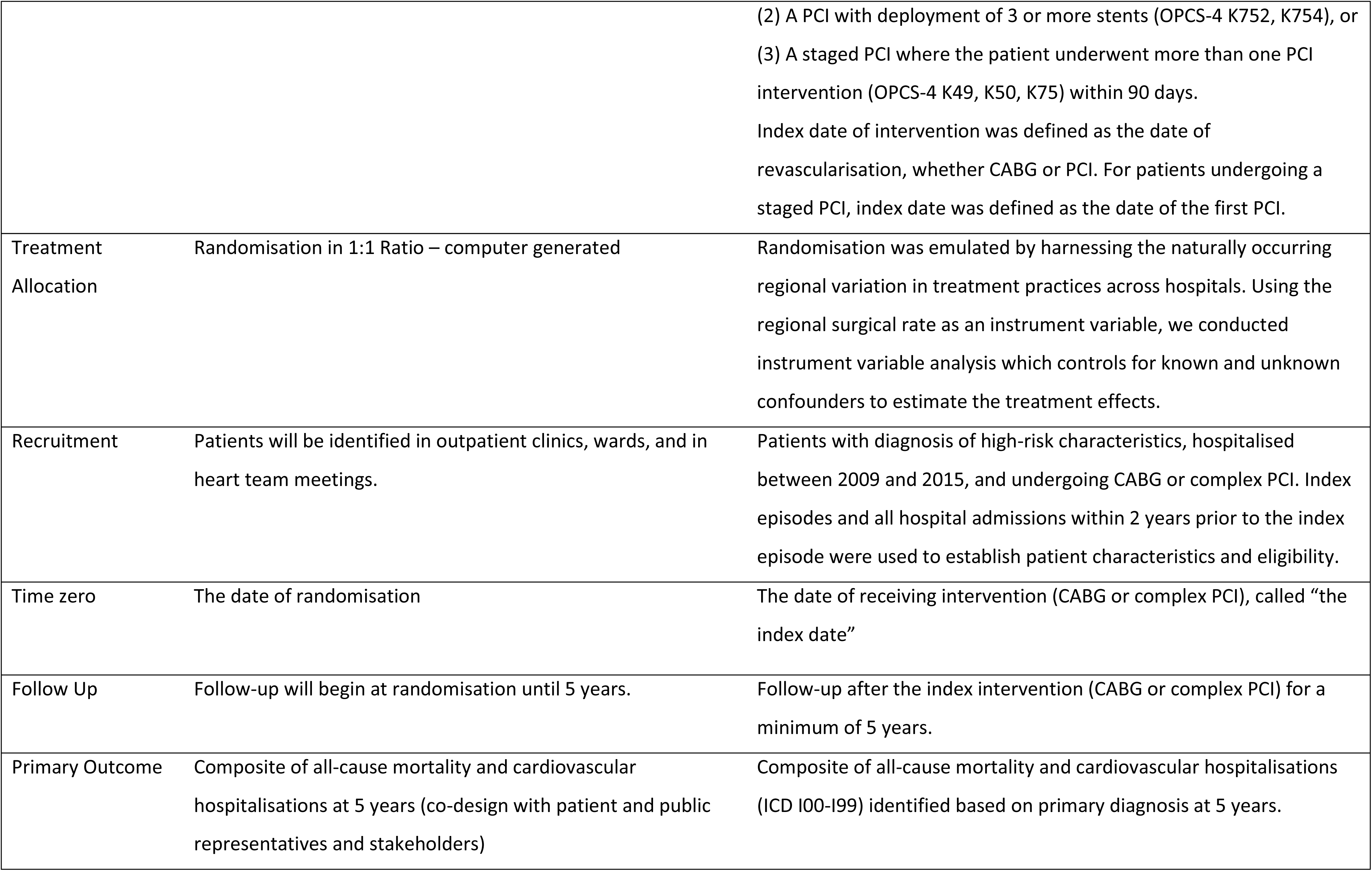
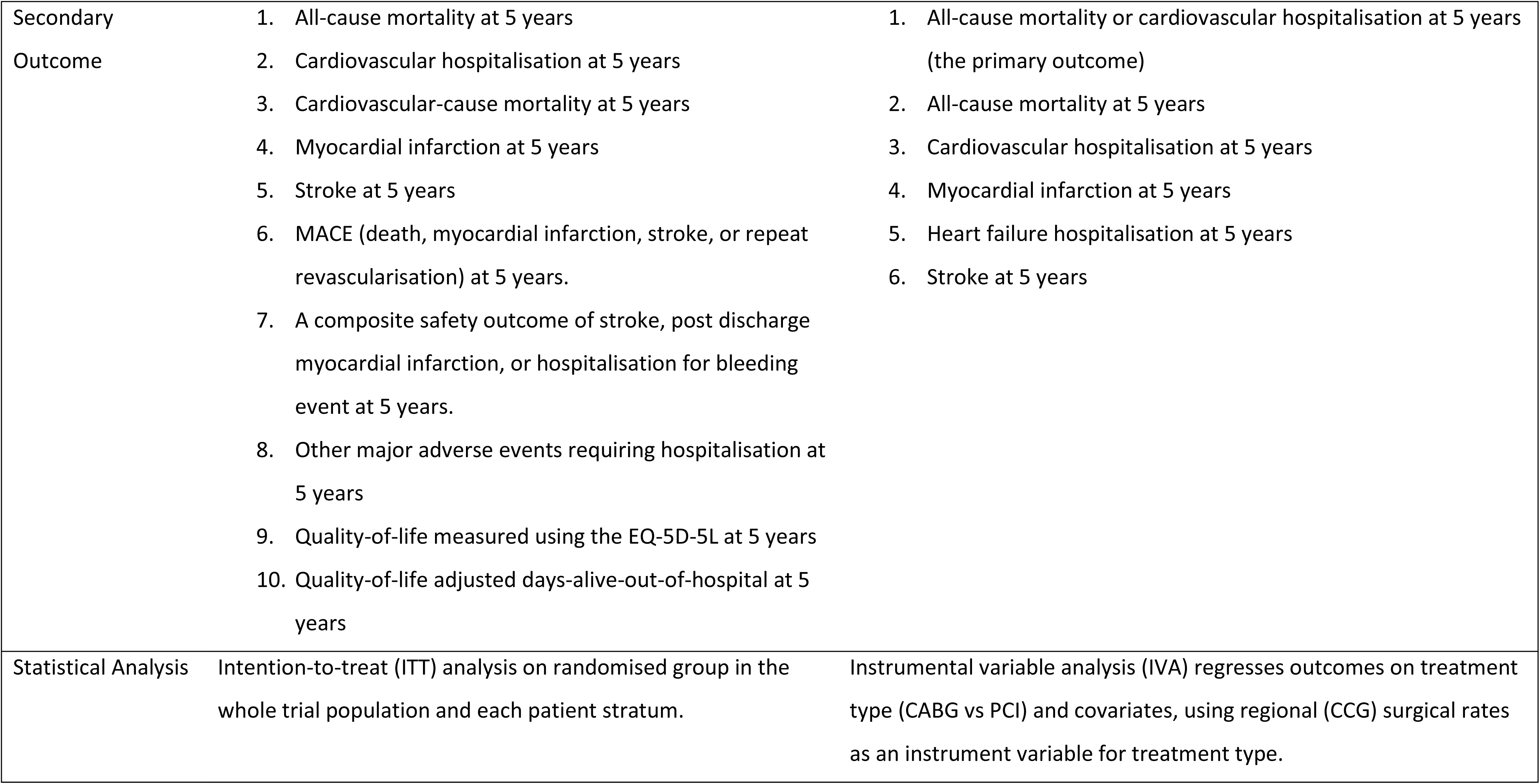
Target trial and emulated trial for a superiority trial of CABG versus PCI in people with high-risk characteristics undergoing coronary revascularisation.

**Figure 1.**
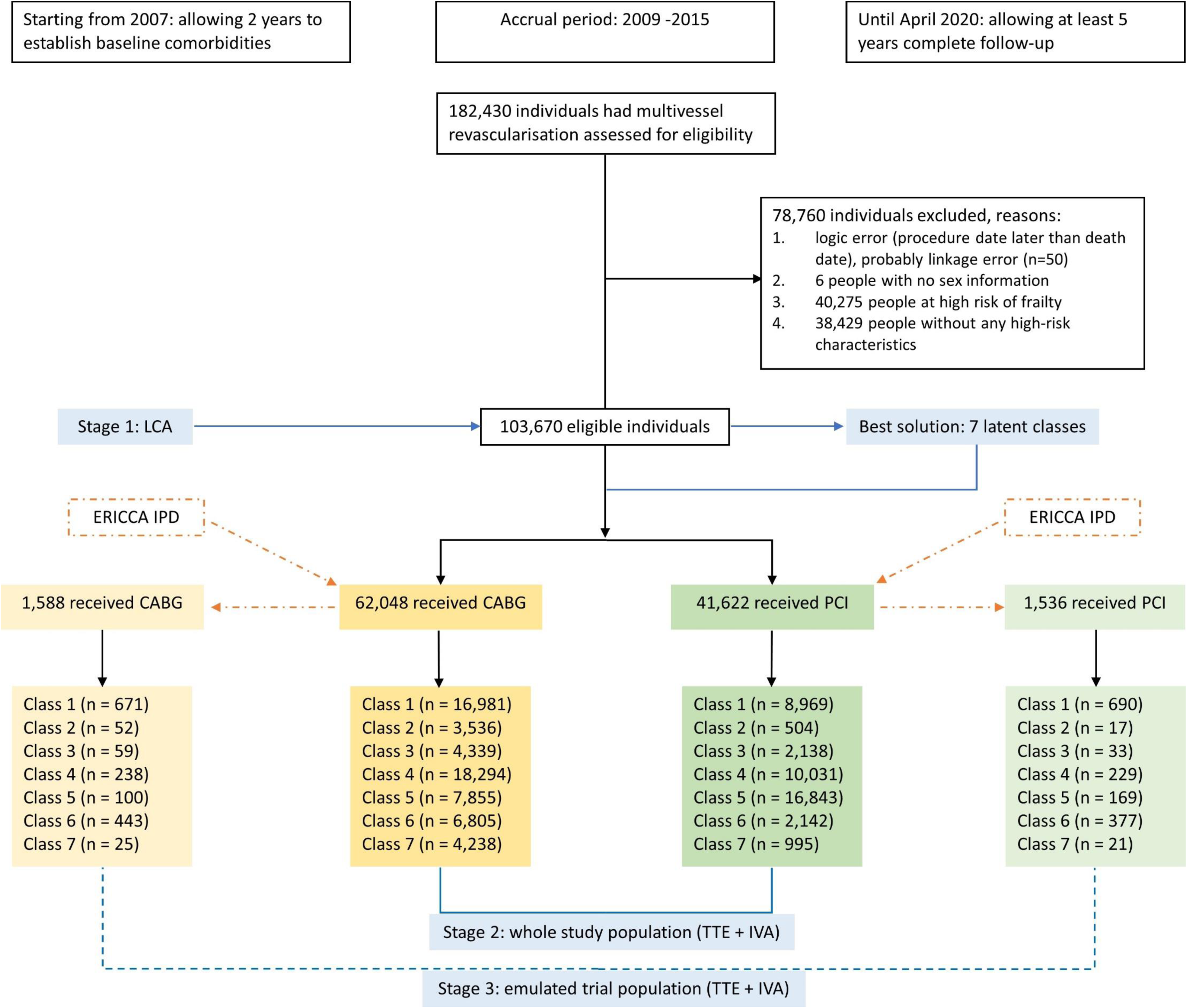
Diagram for participant flow in research stages 1-3. Note: CABG: coronary artery bypass grafting, PCI: percutaneous coronary intervention, LCA: latent class analysis, TTE: target trial emulation, IVA: instrumental variable analysis, ERICCA trial (doi: 10.1056/NEJMoa1413534), IPD: individual participant data. Participant flow is in black. The three research stages are in blue. The dash-dot in orange parts indicate the procedure of propensity score matching (1:1 nearest neighbour) using IPD from the ERICCA trial to match with CABG and PCI separately to derive the CABG and PCI arms in the emulated trial.

Patients’ sociodemographic characteristics, including age, sex, ethnicity, and the index of multiple deprivation (IMD, measuring relative deprivation of patients based on their residential postcodes in England), and clinical characteristics, such as hypertension, ACS, MI, HF, stroke, diabetes, lipidaemia, CKD, PVD, hospital frailty risk score (HFRS), and Charlson comorbidity index were extracted from the HES for further analyses. Multiple imputation with chained equations was used to impute missing data for IMD and ethnicity (10 imputations).

### Stage 1: Latent class analysis (LCA)

LCA^12^ was used to identify patient clusters representing distinctive clinical patterns (phenotypes) from the whole study population (HES). LCA is a probability-based unsupervised learning method. We explored solutions between two and nine latent classes, and determined the optimal number of latent classes for patient stratification based on the following three criteria:

1. Statistical criterion: Akaike Information Criterion (AIC) and Bayesian Information Criterion (BIC) to account for model fit and model complexity. Lower AIC and BIC values indicate better model fit. We also used the elbow method for clustering^13^.
2. Clinical interpretation of latent class patterns: the optimal solution needed to make sense in clinical practice, through discussion with cardiologists and cardiac surgeons in the team. We considered whether new patterns emerged when comparing k with k+1 latent classes, or pattern saturation was reached at k latent classes.
3. Practical considerations to transform the TTE to a two-arm, parallel, multiple strata pragmatic RCT: the number of latent classes and the sample size in each class should be feasible for participant recruitment.

The optimal number of latent classes was decided through consensus of the research team and was used for patient stratification for the following research stages. Class membership for each patient was assigned based on the largest estimated posterior probability from the optimal solution.

### Stage 2: target trial emulation (TTE) for the whole study population (HES)

IVA was used to test the hypothesis that patients in different latent classes representing different phenotypes would have heterogeneous treatment effects (HTE) between CABG and PCI. Previous empirical studies ^14–16^ have shown regional variation in patients receiving CABG or PCI. The standardised CABG rate in each clinical commissioning group (CCG) has been shown as a valid and effective IV ^17,18^, representing random variation in clinician preference and local practice of revascularisation approaches across CCGs. Therefore, we used it as the IV to address both measured and unmeasured confounding, with the two-stage residual inclusion (2SRI) estimator ^19–21^ for time-to-event data to estimate the Local Average Treatment Effects (LATEs) on the composite primary outcome and secondary outcomes for the whole study population (HES) and patients in different latent classes (the **<u>estimand</u>**). LATE is the treatment effect (TE) for people who took the procedure suggested by their care providers (compliers). Since the endogenous variable was binary, the Wald test was used to assess the strength of IV from the first stage using Logistic regression. To recognise statistical uncertainty in the estimates of ATE, bootstrap resampling was performed 1000 times, stratified by CCG, treatment group, death, and censoring status to maintain the structure of the original population and latent classes across replicates^20^, using the ALICE High Performance Computing (HPC) Facility at the University of Leicester, to obtain the standard errors of the 2SRI estimator.^19,20^ We used Cox proportional hazards models for all time-to-event outcomes, and reported the bootstrapped hazard ratios (HR) and 95% bias-corrected and accelerated (BCa) confidence interval (CI) to account for bias and skewness.

### Stage 3: TTE for a likely trial population

Patients participating in RCTs are healthier and carry fewer risk factors than the general patient population (HES) due to the voluntary basis of participation. To obtain better estimates of the ATE in a likely trial population, propensity score matching (PSM) was used to match patients in the HES population to individual participant data (IPD) from the ERICCA trial^22^ that recruited high-risk patients undergoing cardiac surgery in 18 UK centres. Although the research context in the ERICCA trial (evaluating remote ischaemic preconditioning in 1612 heart surgery cases) was different from our target trial, the matching was based on the “overall high-risk” profile of trial participants that are likely to be recruited, using the same infrastructure for cardiac surgery trials across UK centres. We have previously used this approach (PSM) to estimate the treatment effects (TE) in a real-world trial population.^14^ Important baseline covariates available in both ERICCA and HES datasets were used for matching to obtain two arms: ERICCA matched HES CABG (intervention arm) and ERICCA matched HES PCI (control arm) for the TTE. We evaluated whether balance among the covariates was achieved after PSM (1:1 nearest neighbour). The same procedures of IVA with 1000 bootstrap resampling described in Stage 2 were conducted again to estimate the ATE comparing CABG with PCI in the emulated trial population and patients in each latent class for all clinical outcomes, where the class membership was the same as in the whole study population (HES), as the emulated trial population is a subset of all the NHS patients.

### Stage 4: transform to a pragmatic trial

Key issues relating to the trial design including the primary outcome, the number of classes, the desired treatment effect or the minimal clinically important difference (MCID), and the likely attrition and non-adherence to allocation, were discussed at five online international multi-stakeholder coproduction workshops from November 2025 to January 2026.

Because treatment effects (TE) may vary between classes but are likely to be related between classes, we will use Bayesian borrowing, a statistical technique that incorporates external data into a new trial to increase efficiency (particularly in small populations), reduce sample sizes, and lower costs ^23,24^. Eight sets of true class-specific TE were specified to determine the sample size for the target trial using Bayesian borrowing. The global null hypothesis was all TEs are null (i.e. HR=1, or logHR=0); the global alternative hypothesis was all TEs are the target TE, with six partial alternatives as sensitivity analysis (some nulls and some non-nulls, meaning treatment works in some patient classes but not others), including one null (the class with the smallest or largest prevalence) + six non-nulls (2 sets); six nulls + one non-null (the class with the smallest or largest prevalence, 2 sets); three nulls (classes with largest prevalence) + four non-nulls; four nulls + three non-nulls (classes with largest prevalence). In addition, sample sizes for three different trial designs were explored: (1) seven separate trials, one for each class, (2) a single all-comers trial including all high-risk characteristics with subsequent analyses by seven classes, and (3) a single trial stratified by seven classes recruiting participants until the average power for class-specific analysis reached 90%. Overall performance measures included family-wise type 1 error rate (FWER) under the global null and the average power weighted by class prevalence across the non-null classes. Technical details on parameter specification, model assumptions, and computational approach for sample size and power consideration were in the **<u>online supplementary materials – methods</u>**.

## Results

### Study population characteristics

Of 182,430 patients who underwent multivessel revascularisation in England between 2009 and 2015, 103,670 (56.8%) patients had at least one of the seven pre-specified high-risk characteristics and were included as the study population. 62,048 patients (59.9%) received CABG (intervention arm) and 41,622 (40.1%) received PCI (control arm). The median age of the whole study population was 68.5 years (60.2 Q1, 76.0 Q3), with 29,970 (28.9%) >75 years and 27,271 (26.3%) female. The study population was equally distributed in the socioeconomic quintiles, with 2,805 (2.7%) missing. Among them, 84,309 (81.3%) were White, 6,701 (6.5%) Asian, 843 (0.8%) Black, 1,890 (1.8%) mixed/other ethnicities, and 9,927 (9.6%) missing ethnicity. For the high-risk clinical characteristics, 44,809 (43.2%) presented with ACS, 17,154 (16.5%) had a previous heart failure, 8,947 (8.6%) had CKD, 8,038 (7.8%) had PVD, and 49,695 (47.9%) had intermediate risk of frailty.

### Stage 1: LCA results

Model summary statistics from 2 to 9 classes using LCA are in **Table S1** and **Figure S2.** AIC, BIC, G^2^ (likelihood ratio/deviance statistic), and χ^2^ goodness of fit dropped substantially from two to seven classes and stabilised afterwards, with the “elbow” at seven classes (statistical criterion). The frequency and percentage of predicted class memberships from two to nine classes are in **Table S2** (considerations for trial implementation). New patterns continued to emerge from two classes (a class with 100% ACS representation in radar and bar charts, **Figure 2**), then a class with 100% female emerged in the three classes solution, followed by intermediate risk of frailty (four classes), age>75 years (five classes), CKD (six classes), and two new patterns, PVD and HF with 100% representation in each class in the seven classes solution. Patterns were saturated in seven classes, as a repetitive pattern of intermediate frailty emerged in eight classes (class patterns and interpretation). Based on the three criteria, the multidisciplinary research team agreed seven classes were the best solution from LCA. The patterns and feature description of seven classes were summarised in **Figure 3**. The tree structure represented a **<u>stepwise stratification (segmentation)</u>**. The whole study population was first stratified by CKD (Class 6, n=8,947, 8.6%), followed by age>75 years (Class 1, n=29,950, 25.0%), PVD (Class 7, n=5,233, 5.0%), intermediate risk of frailty (Class 4, n=28,325, 27.3%), ACS (Class 5, n=24,698, 23.8%), HF (Class 2, n=4,040, 3.9%), and finally women (Class 3, n=6,477, 6.2%). Patients in each class had that characteristic but no previous characteristics. Patients’ baseline sociodemographic and clinical characteristics by 7 latent classes are in **Table 2**. The proportion of patients received CABG varied among the seven classes, from 31.8% (Class 5) to 87.5% (Class 5), average 59.9% for the whole study population.

**Figure 2.**
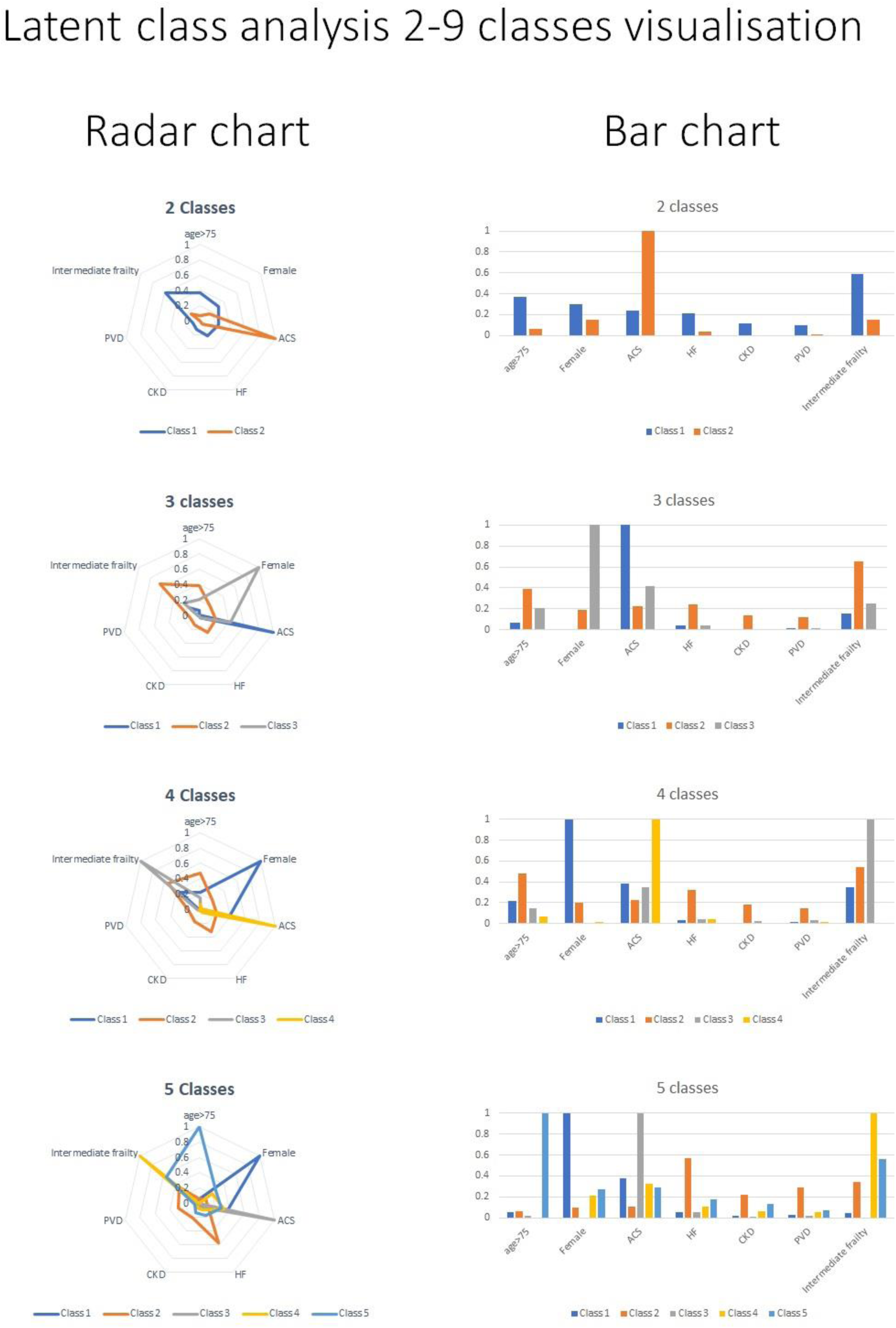
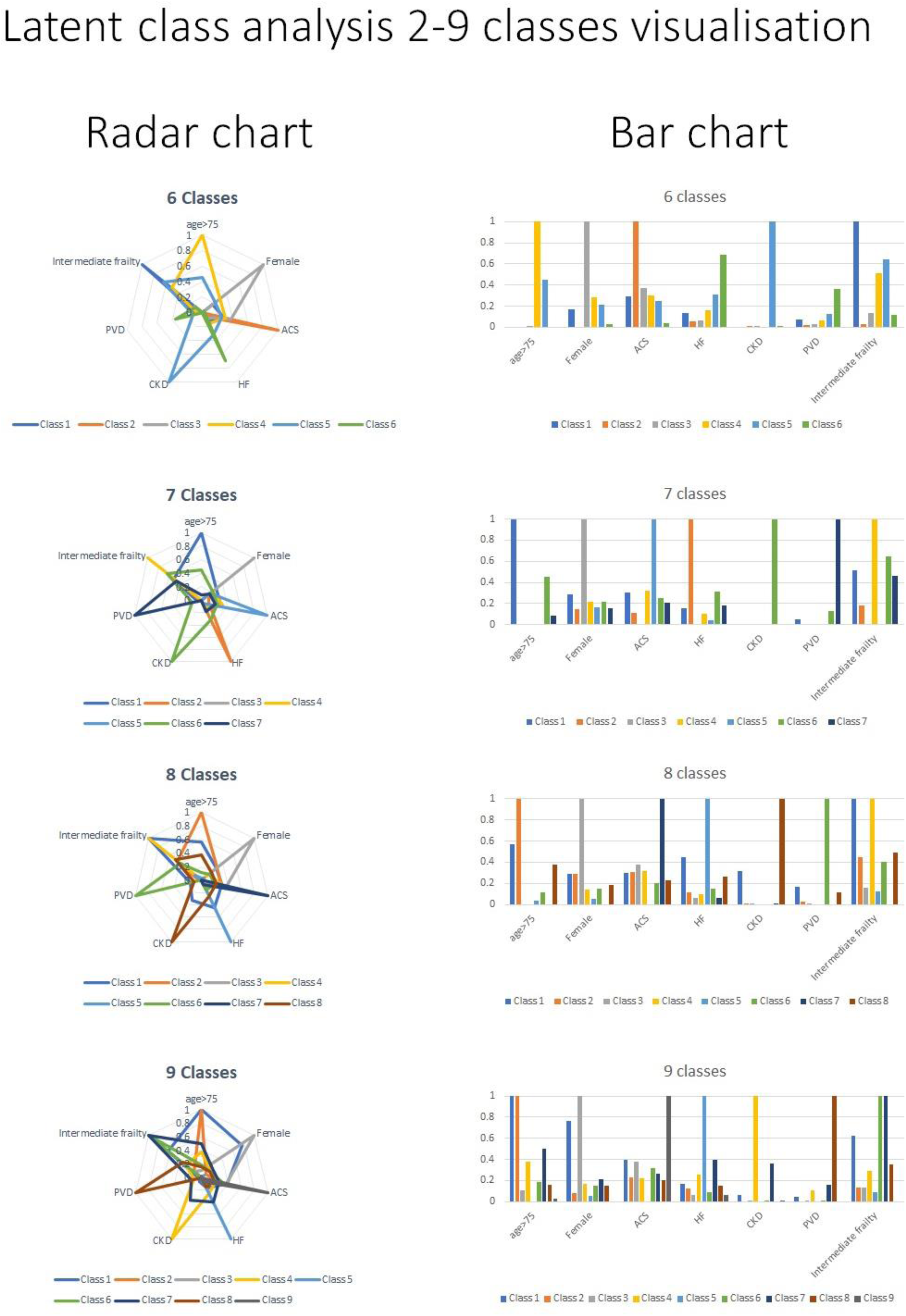
Visualisation of patterns from two to nine classes using latent class analysis (radar and bar charts, stage 1)

**Figure 3.**
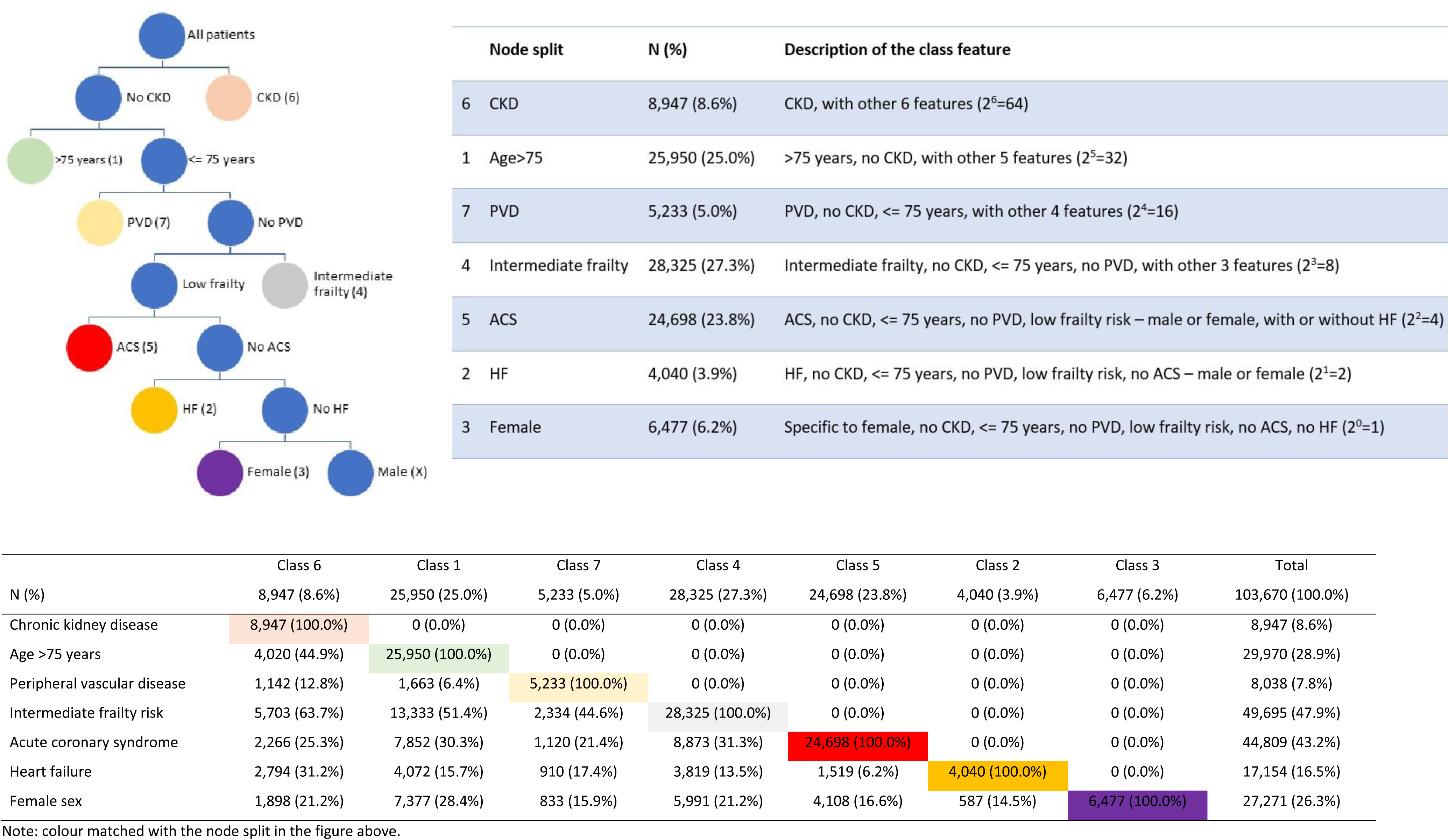
Pattern summary and interpretation of the seven latent classes (optimal solution from latent class analysis, stage 1) Note: colour matched with the node split in the figure above.

**Table 2.** Patients’ baseline sociodemographic and clinical characteristics in the whole study population (HES) by seven latent classes (Stage 2)

|  | Class 6 | Class 1 | Class 7 | Class 4 | Class 5 | Class 2 | Class 3 | Total | P value |
| --- | --- | --- | --- | --- | --- | --- | --- | --- | --- |
| N (%) | 8,947 (8.6%) | 25,950 (25.0%) | 5,233 (5.0%) | 28,325 (27.3%) | 24,698 (23.8%) | 4,040 (3.9%) | 6,477 (6.2%) | 103,670 (100.0%) |  |
| Revascularisation approach |  |  |  |  |  |  |  |  |  |
| PCI | 2,142 (23.9%) | 8,969 (34.6%) | 995 (19.0%) | 10,031 (35.4%) | 16,843 (68.2%) | 504 (12.5%) | 2,138 (33.0%) | 41,622 (40.1%) | <0.001 |
| CABG | 6,805 (76.1%) | 16,981 (65.4%) | 4,238 (81.0%) | 18,294 (64.6%) | 7,855 (31.8%) | 3,536 (87.5%) | 4,339 (67.0%) | 62,048 (59.9%) |  |
| Age | 72.5 (9.2) | 79.6 (3.5) | 64.2 (7.3) | 64.0 (8.1) | 59.6 (9.0) | 62.7 (8.3) | 63.2 (8.4) | 67.5 (10.8) | <0.001 |
| ≤75 years | 4,927 (55.1%) | 0 (0.0%) | 5,233 (100.0%) | 28,325 (100.0%) | 24,698 (100.0%) | 4,040 (100.0%) | 6,477 (100.0%) | 73,700 (71.1%) | <0.001 |
| >75 years | 4,020 (44.9%) | 25,950 (100.0%) | 0 (0.0%) | 0 (0.0%) | 0 (0.0%) | 0 (0.0%) | 0 (0.0%) | 29,970 (28.9%) |  |
| Sex |  |  |  |  |  |  |  |  |  |
| Male | 7,049 (78.8%) | 18,573 (71.6%) | 4,400 (84.1%) | 22,334 (78.8%) | 20,590 (83.4%) | 3,453 (85.5%) | 0 (0.0%) | 76,399 (73.7%) | <0.001 |
| Female | 1,898 (21.2%) | 7,377 (28.4%) | 833 (15.9%) | 5,991 (21.2%) | 4,108 (16.6%) | 587 (14.5%) | 6,477 (100.0%) | 27,271 (26.3%) |  |
| Ethnicity |  |  |  |  |  |  |  |  |  |
| White | 7,216 (87.9%) | 21,965 (93.8%) | 4,520 (94.1%) | 22,886 (88.6%) | 19,389 (88.0%) | 3,309 (89.4%) | 5,024 (87.6%) | 84,309 (89.9%) | <0.001 |
| Asian | 743 (9.0%) | 978 (4.2%) | 190 (4.0%) | 2,164 (8.4%) | 1,860 (8.4%) | 274 (7.4%) | 492 (8.6%) | 6,701 (7.1%) |  |
| Black | 102 (1.2%) | 145 (0.6%) | 30 (0.6%) | 250 (1.0%) | 198 (0.9%) | 38 (1.0%) | 80 (1.4%) | 843 (0.9%) |  |
| Mixed/others | 152 (1.9%) | 337 (1.4%) | 61 (1.3%) | 543 (2.1%) | 582 (2.6%) | 79 (2.1%) | 136 (2.4%) | 1,890 (2.0%) |  |
| Index of Multiple Deprivation score (continuous) | 20.7 (14.8) | 18.0 (13.5) | 24.8 (16.8) | 23.0 (15.9) | 22.0 (15.6) | 21.8 (15.2) | 21.9 (15.4) | 21.3 (15.3) | <0.001 |
| Index of Multiple Deprivation (quintile) |  |  |  |  |  |  |  |  |  |
| Least deprived quintile (0-20%) | 1,701 (19.0%) | 6,399 (24.7%) | 744 (14.2%) | 4,762 (16.8%) | 4,496 (18.2%) | 702 (17.4%) | 1,181 (18.2%) | 19,985 (19.3%) | <0.001 |
| Less deprived quintile | 1,870 (20.9%) | 6,048 (23.3%) | 887 (17.0%) | 5,373 (19.0%) | 4,775 (19.3%) | 816 (20.2%) | 1,211 (18.7%) | 20,980 (20.2%) |  |
| Middle quintile | 1,782 (19.9%) | 4,425 (17.1%) | 1,055 (20.2%) | 5,690 (20.1%) | 4,819 (19.5%) | 822 (20.3%) | 1,272 (19.6%) | 19,865 (19.2%) |  |
| More deprived quintile | 1,683 (18.8%) | 4,143 (16.0%) | 1,101 (21.0%) | 6,044 (21.3%) | 4,862 (19.7%) | 793 (19.6%) | 1,290 (19.9%) | 19,916 (19.2%) |  |
| Most deprived quintile | 1,625 (18.2%) | 4,078 (15.7%) | 1,236 (23.6%) | 6,089 (21.5%) | 5,038 (20.4%) | 787 (19.5%) | 1,266 (19.5%) | 20,119 (19.4%) |  |
| Unknown | 286 (3.2%) | 857 (3.3%) | 210 (4.0%) | 367 (1.3%) | 708 (2.9%) | 120 (3.0%) | 257 (4.0%) | 2,805 (2.7%) |  |
| Hypertension |  |  |  |  |  |  |  |  |  |
| No | 1,074 (12.0%) | 8,267 (31.9%) | 1,036 (19.8%) | 9,741 (34.4%) | 15,365 (62.2%) | 1,250 (30.9%) | 2,284 (35.3%) | 39,017 (37.6%) | <0.001 |
| Yes | 7,873 (88.0%) | 17,683 (68.1%) | 4,197 (80.2%) | 18,584 (65.6%) | 9,333 (37.8%) | 2,790 (69.1%) | 4,193 (64.7%) | 64,653 (62.4%) |  |
| Myocardial infarction |  |  |  |  |  |  |  |  |  |
| No | 5,639 (63.0%) | 18,338 (70.7%) | 3,595 (68.7%) | 19,316 (68.2%) | 12,227 (49.5%) | 2,728 (67.5%) | 5,500 (84.9%) | 67,343 (65.0%) | <0.001 |
| Yes | 3,308 (37.0%) | 7,612 (29.3%) | 1,638 (31.3%) | 9,009 (31.8%) | 12,471 (50.5%) | 1,312 (32.5%) | 977 (15.1%) | 36,327 (35.0%) |  |
| Acute Coronary Syndrome |  |  |  |  |  |  |  |  |  |
| No | 6,681 (74.7%) | 18,098 (69.7%) | 4,113 (78.6%) | 19,452 (68.7%) | 0 (0.0%) | 4,040 (100.0%) | 6,477 (100.0%) | 58,861 (56.8%) | <0.001 |
| Yes | 2,266 (25.3%) | 7,852 (30.3%) | 1,120 (21.4%) | 8,873 (31.3%) | 24,698 (100.0%) | 0 (0.0%) | 0 (0.0%) | 44,809 (43.2%) |  |
| Heart failure |  |  |  |  |  |  |  |  |  |
| No | 6,153 (68.8%) | 21,878 (84.3%) | 4,323 (82.6%) | 24,506 (86.5%) | 23,179 (93.8%) | 0 (0.0%) | 6,477 (100.0%) | 86,516 (83.5%) | <0.001 |
| Yes | 2,794 (31.2%) | 4,072 (15.7%) | 910 (17.4%) | 3,819 (13.5%) | 1,519 (6.2%) | 4,040 (100.0%) | 0 (0.0%) | 17,154 (16.5%) |  |
| Chronic kidney disease |  |  |  |  |  |  |  |  |  |
| No | 0 (0.0%) | 25,950 (100.0%) | 5,233 (100.0%) | 28,325 (100.0%) | 24,698 (100.0%) | 4,040 (100.0%) | 6,477 (100.0%) | 94,723 (91.4%) | <0.001 |
| Yes | 8,947 (100.0%) | 0 (0.0%) | 0 (0.0%) | 0 (0.0%) | 0 (0.0%) | 0 (0.0%) | 0 (0.0%) | 8,947 (8.6%) |  |
| Peripheral vascular disease |  |  |  |  |  |  |  |  |  |
| No | 7,805 (87.2%) | 24,287 (93.6%) | 0 (0.0%) | 28,325 (100.0%) | 24,698 (100.0%) | 4,040 (100.0%) | 6,477 (100.0%) | 95,632 (92.2%) | <0.001 |
| Yes | 1,142 (12.8%) | 1,663 (6.4%) | 5,233 (100.0%) | 0 (0.0%) | 0 (0.0%) | 0 (0.0%) | 0 (0.0%) | 8,038 (7.8%) |  |
| Diabetes |  |  |  |  |  |  |  |  |  |
| No | 5,284 (59.1%) | 20,836 (80.3%) | 3,483 (66.6%) | 20,532 (72.5%) | 21,586 (87.4%) | 3,055 (75.6%) | 5,085 (78.5%) | 79,861 (77.0%) | <0.001 |
| Yes | 3,663 (40.9%) | 5,114 (19.7%) | 1,750 (33.4%) | 7,793 (27.5%) | 3,112 (12.6%) | 985 (24.4%) | 1,392 (21.5%) | 23,809 (23.0%) |  |
| Cerebrovascular disease |  |  |  |  |  |  |  |  |  |
| No | 8,285 (92.6%) | 24,730 (95.3%) | 4,747 (90.7%) | 27,112 (95.7%) | 24,390 (98.8%) | 3,911 (96.8%) | 6,334 (97.8%) | 99,509 (96.0%) | <0.001 |
| Yes | 662 (7.4%) | 1,220 (4.7%) | 486 (9.3%) | 1,213 (4.3%) | 308 (1.2%) | 129 (3.2%) | 143 (2.2%) | 4,161 (4.0%) |  |
| Lipidaemia |  |  |  |  |  |  |  |  |  |
| No | 3,223 (36.0%) | 12,603 (48.6%) | 1,290 (24.7%) | 12,170 (43.0%) | 16,316 (66.1%) | 1,416 (35.0%) | 2,465 (38.1%) | 49,483 (47.7%) | <0.001 |
| Yes | 5,724 (64.0%) | 13,347 (51.4%) | 3,943 (75.3%) | 16,155 (57.0%) | 8,382 (33.9%) | 2,624 (65.0%) | 4,012 (61.9%) | 54,187 (52.3%) |  |
| Hospital frailty risk scores (continuous) | 7.1 (4.3) | 5.8 (4.4) | 5.2 (4.3) | 8.8 (2.8) | 1.4 (1.5) | 1.8 (1.6) | 1.6 (1.5) | 5.2 (4.4) | <0.001 |
| Frailty risk (categorical) |  |  |  |  |  |  |  |  |  |
| Low frailty risk | 3,244 (36.3%) | 12,617 (48.6%) | 2,899 (55.4%) | 0 (0.0%) | 24,698 (100.0%) | 4,040 (100.0%) | 6,477 (100.0%) | 53,975 (52.1%) | <0.001 |
| Intermediate frailty risk | 5,703 (63.7%) | 13,333 (51.4%) | 2,334 (44.6%) | 28,325 (100.0%) | 0 (0.0%) | 0 (0.0%) | 0 (0.0%) | 49,695 (47.9%) |  |
| Charlson Comorbidity Index (continuous) | 8.6 (6.0) | 4.8 (4.8) | 6.9 (5.6) | 6.3 (5.9) | 3.2 (3.2) | 3.9 (3.2) | 2.4 (3.3) | 5.1 (5.2) | <0.001 |

### Stage 2: TTE results for the whole study population

The crude event rates for the composite primary outcome and secondary outcomes between CABG and PCI over 5 years of follow-up among the seven classes are in **Table S3**. Regional variation in the standardised CABG rates by CCG in England and the strength as an instrumental variable (IV), overall and for each class, are in **Figure S4** and **Table S4,** respectively. The standardised CABG rates by CCG was a valid IV. Hazard ratios with 95% BCa-CI for the composite primary outcome and secondary outcomes in the whole study population and the seven classes are in **Table 3 (upper panel)**. The results were colour-coded, favouring CABG highlighted in light blue in the cell, favouring PCI in light orange, and no difference (95% BCaCI cross 1) with no highlight. Compared with PCI, CABG had a lower event rate for the composite primary outcome in the whole study population 0.46 (0.43 to 0.49) and six classes, with effect sizes ranging from 0.27 (0.22 to 0.32) in Class 5 to 0.76 (0.64 to 0.91) in Class 6, except for Class 2 favouring PCI 2.77 (1.43 to 4.76).

**Table 3.** Hazard ratios with 95% bias-corrected and accelerated (BCa) confidence interval (CI) using instrumental variable analysis for the composite primary and secondary outcomes at 5 years in the whole study population (HES data), the emulated trial (ERICCA matched HES data) and the seven classes (Stages 2 and 3)

|  | All-cause mortality or<br>cardiovascular<br>hospitalisation | All-cause mortality | Cardiovascular mortality | Cardiovascular<br>rehospitalisation | Myocardial infarction | Heart failure<br>hospitalisation | Stroke |
| --- | --- | --- | --- | --- | --- | --- | --- |
| All (HES) | 0.46 (0.43 to 0.49) | 0.84 (0.73 to 0.97) | 0.78 (0.68 to 0.91) | 0.37 (0.35 to 0.40) | 0.39 (0.34 to 0.46) | 0.77 (0.65 to 0.91) | 0.60 (0.49 to 0.73) |
| Class 1 | 0.44 (0.40 to 0.48) | 0.60 (0.50 to 0.69) | 0.58 (0.48 to 0.71) | 0.36 (0.32 to 0.41) | 0.39 (0.31 to 0.47) | 0.54 (0.44 to 0.67) | 0.37 (0.29 to 0.46) |
| Class 2 | 2.77 (1.43 to 4.76) | 11.98 (2.91 to 40.24) | 8.08 (1.50 to 27.27) | 1.37 (0.59 to 2.45) | 4.08 (0.92 to 10.00) | 2.57 (0.83 to 5.76) | 0.13 (0.03 to 0.32) |
| Class 3 | 0.32 (0.26 to 0.38) | 1.01 (0.33 to 2.22) | 0.81 (0.23 to 2.01) | 0.25 (0.20 to 0.31) | 0.44 (0.27 to 0.64) | 1.54 (1.01 to 2.15) | 4.59 (2.50 to 6.67) |
| Class 4 | 0.53 (0.47 to 0.59) | 1.29 (0.91 to 1.87) | 1.09 (0.72 to 1.55) | 0.47 (0.41 to 0.53) | 0.42 (0.31 to 0.53) | 1.11 (0.85 to 1.52) | 0.82 (0.61 to 1.11) |
| Class 5 | 0.27 (0.22 to 0.32) | 0.97 (0.56 to 1.47) | 1.49 (0.87 to 2.41) | 0.21 (0.18 to 0.26) | 0.28 (0.21 to 0.39) | 0.67 (0.37 to 1.12) | 0.73 (0.45 to 1.07) |
| Class 6 | 0.76 (0.64 to 0.91) | 0.85 (0.63 to 1.14) | 0.85 (0.59 to 1.13) | 0.61 (0.46 to 0.78) | 0.25 (0.18 to 0.35) | 0.65 (0.45 to 0.92) | 0.50 (0.32 to 0.76) |
| Class 7 | 0.41 (0.33 to 0.50) | 0.47 (0.26 to 0.78) | 0.40 (0.21 to 0.76) | 0.43 (0.34 to 0.57) | 0.59 (0.38 to 0.89) | 0.94 (0.55 to 1.51) | 0.63 (0.35 to 1.05) |
| Emulated (ERICCA<br>matched HES) | 0.55 (0.46 to 0.65) | 0.85 (0.53 to 1.26) | 0.86 (0.50 to 1.36) | 0.37 (0.27 to 0.48) | 0.34 (0.25 to 0.47) | 1.22 (0.84 to 1.79) | 0.86 (0.57 to 1.22) |
| Class 1 | 0.54 (0.44 to 0.65) | 0.49 (0.31 to 0.79) | 0.45 (0.27 to 0.78) | 0.37 (0.28 to 0.54) | 0.65 (0.38 to 0.99) | 0.50 (0.33 to 0.74) | 0.62 (0.36 to 1.00) |
| Class 2 | Unstable estimation | Unstable estimation | Unstable estimation | Unstable estimation | Not able to estimate | Unstable estimation | Not able to estimate |
| Class 3 | 0.14 (0.09 to 0.20) | 0.76 (0.31 to 2.56) | 0.32 (0.02 to 1.08) | 0.13 (0.08 to 0.18) | Not able to estimate | Unstable estimation | Not able to estimate |
| Class 4 | 0.64 (0.43 to 0.95) | 4.55 (1.02 to 12.40) | 2.01 (0.32 to 6.52) | 0.55 (0.29 to 0.91) | 5.80 (2.02 to 17.33) | 0.92 (0.07 to 2.23) | Unstable estimation |
| Class 5 | 0.74 (0.46 to 1.14) | 12.74 (0.11 to 54.08) | 25.16 (1.66 to 77.47) | 0.67 (0.40 to 1.07) | 0.06 (0.02 to 0.11) | Unstable estimation | Not able to estimate |
| Class 6 | 0.66 (0.43 to 1.01) | 2.68 (1.07 to 5.02) | 2.37 (0.90 to 5.23) | 0.30 (0.13 to 0.61) | 0.12 (0.04 to 0.31) | 7.75 (1.60 to 17.87) | 0.79 (0.19 to 1.78) |
| Class 7 | 0.79 (0.21 to 1.50) | 0.38 (0.05 to 1.13) | 0.45 (0.12 to 1.02) | 0.41 (0.01 to 2.07) | Unstable estimation | Not able to estimate | Not able to estimate |

Note: the hazard ratios and 95% bias-corrected and accelerated (BCa) confidence intervals (CI) to account for both bias and skewness in the table above were based on 1000 bootstrap replications for each calculation.
Results were colour-coded, favouring CABG highlighted in light blue in the cell, favouring PCI in light orange, no difference (95% BCa CI cross 1) with no highlight.

The ATEs favoured CABG, consistent across all secondary outcomes for the whole study population. All-cause mortality and cardiovascular mortality shared the same patterns among the seven classes, with results favouring CABG in Classes 1 and 7, favouring PCI in Class 2 (although wide CI), and no differences in Classes 3, 4, 5, and 6. CABG had better outcomes for cardiovascular hospitalisation and MI in six classes, except for Class 2 (no difference). For heart failure hospitalisation, results favoured CABG in Classes 1 and 6, favouring PCI in Class 3, and no differences in Classes 2, 4, 5, and 7. For stroke, results favoured CABG in Classes 1, 2, and 6, favouring PCI in Class 3, and no differences in Classes 4, 5, and 7 (**Table 3** upper panel, by column).

Only Class 1 had consistent results favouring CABG across all clinical outcomes. Treatment effects favoured CABG or PCI were inconsistent among different clinical outcomes in Classes 2 and 3. Classes 2 favoured PCI in the composite primary outcome, mainly due to mortality. However, patients underwent CABG in Class 2 had fewer numbers of strokes. Class 3 had favourable outcomes for CABG in the composite primary outcome, cardiovascular hospitalisation, and MI, but not heart failure hospitalisation and stroke (favoured PCI). For Classes 4, 5, 6, and 7, the results favoured CABG in the composite primary outcome and some secondary outcomes, with other secondary outcomes showing no difference from PCI (**Table 3** upper panel, by row).

### Stage 3: TTE results for a likely trial population

PSM with 1:1 nearest neighbour matching derived 3,124 participants for the emulated trial, with 1,588 (50.8%) in the CABG arm and 1,536 (49.2%) in the PCI arm. Variables before and after PSM between the ERICCA trial and the HES CABG and PCI arms for the emulated trial are in **Tables S5** and **S6,** and illustrated in **Figure S6**. Participants in the ERICCA trial were older (76.2±6.5 years) than patients received CABG (68.8±10.0 years) and PCI (65.5±11.7 years) in the complete HES data; a greater proportion of White people, had a higher comorbidity burden in CKD, HF, lipidaemia, cerebrovascular accident (CVA/stroke) and MI than both CABG and PCI cohorts in HES (**Table 4, Figure S6**). Balance of variables was achieved after PSM, as the standardised percentages of bias were around 0 (**Figure S6**).

**Table 4.** Baseline sociodemographic and clinical characteristics in the whole study population (HES), the ERICCA trial, and the emulated trial (ERICCA matched HES data, Stages 2 and 3)

|  | HES | ERICCA Trial | Emulated (ERICCA matched HES) | The whole HES population |  |  | Emulated (ERICCA matched HES) |  |  |
| --- | --- | --- | --- | --- | --- | --- | --- | --- | --- |
|  | Total |  | Total | CABG | PCI | P value | CABG | PCI | P value |
| N (%) | 103,670 | 1,596 | 3,124 | 62,048 (59.9%) | 41,622 (40.1%) |  | 1,588 (50.8%) | 1,536 (49.2%) |  |
| Age | 67.5 (10.8) | 76.2 (6.5) | 76.4 (7.7) | 68.8 (10.0) | 65.5 (11.7) | <0.001 | 76.2 (7.0) | 76.7 (8.3) | 0.063 |
| ≤75 years | 73,700 (71.1%) | 585 (36.7%) | 1,196 (38.3%) | 31,564 (75.8%) | 42,136 (67.9%) | <0.001 | 586 (38.2%) | 610 (38.4%) | 0.880 |
| >75 years | 29,970 (28.9%) | 1,011 (63.3%) | 1,928 (61.7%) | 10,058 (24.2%) | 19,912 (32.1%) |  | 950 (61.8%) | 978 (61.6%) |  |
| Sex |  |  |  |  |  |  |  |  |  |
| Male | 76,399 (73.7%) | 1,142 (71.6%) | 2,228 (71.3%) | 45,980 (74.1%) | 30,419 (73.1%) | <0.001 | 1,115 (70.2%) | 1,113 (72.5%) | 0.165 |
| Female | 27,271 (26.3%) | 454 (28.4%) | 896 (28.7%) | 16,068 (25.9%) | 11,203 (26.9%) |  | 473 (29.8%) | 423 (27.5%) |  |
| Ethnicity |  |  |  |  |  |  |  |  |  |
| White | 84,309 (89.9%) | 1,506 (94.4%) | 2,699 (94.5%) | 50,872 (90.3%) | 33,437 (89.4%) | <0.001 | 1,377 (94.8%) | 1,322 (94.1%) | 0.330 |
| Asian | 6,701 (7.1%) | 77 (4.8%) | 136 (4.8%) | 3,917 (7.0%) | 2,784 (7.4%) |  | 61 (4.2%) | 75 (5.3%) |  |
| Black | 843 (0.9%) | 8 (0.5%) | 11 (0.4%) | 455 (0.8%) | 388 (1.0%) |  | 7 (0.5%) | 4 (0.3%) |  |
| Mixed/others | 1,890 (2.0%) | 5 (0.3%) | 11 (0.4%) | 1,098 (1.9%) | 792 (2.1%) |  | 7 (0.5%) | 4 (0.3%) |  |
| Index of Multiple Deprivation score (continuous) | 21.3 (15.3) |  | 19.3 (14.5) | 21.0 (15.1) | 21.7 (15.5) | <0.001 | 18.7 (14.1) | 20.0 (14.8) | 0.014 |
| Index of Multiple Deprivation (quintile) |  |  |  |  |  |  |  |  |  |
| Least deprived quintile (0-20%) | 19,985 (19.3%) |  | 735 (23.5%) | 12,133 (19.6%) | 7,852 (18.9%) | <0.001 | 387 (24.4%) | 348 (22.7%) | 0.025 |
| Less deprived quintile | 20,980 (20.2%) |  | 646 (20.7%) | 12,739 (20.5%) | 8,241 (19.8%) |  | 342 (21.5%) | 304 (19.8%) |  |
| Middle quintile | 19,865 (19.2%) |  | 550 (17.6%) | 11,606 (18.7%) | 8,259 (19.8%) |  | 258 (16.2%) | 292 (19.0%) |  |
| More deprived quintile | 19,916 (19.2%) |  | 545 (17.4%) | 11,690 (18.8%) | 8,226 (19.8%) |  | 273 (17.2%) | 272 (17.7%) |  |
| Most deprived quintile | 20,119 (19.4%) |  | 571 (18.3%) | 11,851 (19.1%) | 8,268 (19.9%) |  | 278 (17.5%) | 293 (19.1%) |  |
| Unknown | 2,805 (2.7%) |  | 77 (2.5%) | 2,029 (3.3%) | 776 (1.9%) |  | 50 (3.1%) | 27 (1.8%) |  |
| Hypertension |  |  |  |  |  |  |  |  |  |
| No | 39,017 (37.6%) | 395 (24.7%) | 734 (23.5%) | 14,914 (24.0%) | 24,103 (57.9%) | <0.001 | 383 (24.1%) | 351 (22.9%) | 0.404 |
| Yes | 64,653 (62.4%) | 1,201 (75.3%) | 2,390 (76.5%) | 47,134 (76.0%) | 17,519 (42.1%) |  | 1,205 (75.9%) | 1,185 (77.1%) |  |
| Myocardial infarction |  |  |  |  |  |  |  |  |  |
| No | 67,343 (65.0%) | 959 (60.1%) | 1,854 (59.3%) | 40,011 (64.5%) | 27,332 (65.7%) | <0.001 | 973 (61.3%) | 881 (57.4%) | 0.026 |
| Yes | 36,327 (35.0%) | 637 (39.9%) | 1,270 (40.7%) | 22,037 (35.5%) | 14,290 (34.3%) |  | 615 (38.7%) | 655 (42.6%) |  |
| Acute Coronary Syndrome |  |  |  |  |  |  |  |  |  |
| No | 58,861 (56.8%) |  | 1,985 (63.5%) | 13,217 (31.8%) | 45,644 (73.6%) | <0.001 | 765 (49.8%) | 1,220 (76.8%) | <0.001 |
| Yes | 44,809 (43.2%) |  | 1,139 (36.5%) | 28,405 (68.2%) | 16,404 (26.4%) |  | 771 (50.2%) | 368 (23.2%) |  |
| Heart failure |  |  |  |  |  |  |  |  |  |
| No | 86,516 (83.5%) | 1,118 (70.1%) | 2,209 (70.7%) | 48,311 (77.9%) | 38,205 (91.8%) | <0.001 | 1,098 (69.1%) | 1,111 (72.3%) | 0.050 |
| Yes | 17,154 (16.5%) | 478 (29.9%) | 915 (29.3%) | 13,737 (22.1%) | 3,417 (8.2%) |  | 490 (30.9%) | 425 (27.7%) |  |
| Chronic kidney disease |  |  |  |  |  |  |  |  |  |
| No | 94,723 (91.4%) | 1,141 (71.5%) | 2,304 (73.8%) | 55,243 (89.0%) | 39,480 (94.9%) | <0.001 | 1,145 (72.1%) | 1,159 (75.5%) | 0.033 |
| Yes | 8,947 (8.6%) | 455 (28.5%) | 820 (26.2%) | 6,805 (11.0%) | 2,142 (5.1%) |  | 443 (27.9%) | 377 (24.5%) |  |
| Peripheral vascular disease |  |  |  |  |  |  |  |  |  |
| No | 95,632 (92.2%) | 1,475 (92.4%) | 2,908 (93.1%) | 55,654 (89.7%) | 39,978 (96.1%) | <0.001 | 1,483 (93.4%) | 1,425 (92.8%) | 0.499 |
| Yes | 8,038 (7.8%) | 121 (7.6%) | 216 (6.9%) | 6,394 (10.3%) | 1,644 (3.9%) |  | 105 (6.6%) | 111 (7.2%) |  |
| Diabetes |  |  |  |  |  |  |  |  |  |
| No | 79,861 (77.0%) | 1,182 (74.1%) | 2,337 (74.8%) | 45,020 (72.6%) | 34,841 (83.7%) | <0.001 | 1,193 (75.1%) | 1,144 (74.5%) | 0.677 |
| Yes | 23,809 (23.0%) | 414 (25.9%) | 787 (25.2%) | 17,028 (27.4%) | 6,781 (16.3%) |  | 395 (24.9%) | 392 (25.5%) |  |
| Cerebrovascular disease |  |  |  |  |  |  |  |  |  |
| No | 99,509 (96.0%) | 1,412 (88.5%) | 2,839 (90.9%) | 58,434 (94.2%) | 41,075 (98.7%) | <0.001 | 1,411 (88.9%) | 1,428 (93.0%) | <0.001 |
| Yes | 4,161 (4.0%) | 184 (11.5%) | 285 (9.1%) | 3,614 (5.8%) | 547 (1.3%) |  | 177 (11.1%) | 108 (7.0%) |  |
| Lipidaemia |  |  |  |  |  |  |  |  |  |
| No | 49,483 (47.7%) | 472 (29.6%) | 924 (29.6%) | 20,270 (32.7%) | 29,213 (70.2%) | <0.001 | 460 (29.0%) | 464 (30.2%) | 0.447 |
| Yes | 54,187 (52.3%) | 1,124 (70.4%) | 2,200 (70.4%) | 41,778 (67.3%) | 12,409 (29.8%) |  | 1,128 (71.0%) | 1,072 (69.8%) |  |
| Hospital frailty risk scores (continuous) | 5.2 (4.4) |  | 6.4 (4.5) | 5.8 (4.3) | 4.5 (4.3) | <0.001 | 6.3 (4.4) | 6.5 (4.6) | 0.179 |
| Frailty risk (categorical) |  |  |  |  |  |  |  |  |  |
| Low frailty risk | 53,975 (52.1%) |  | 1,300 (41.6%) | 28,694 (46.2%) | 25,281 (60.7%) | <0.001 | 676 (42.6%) | 624 (40.6%) | 0.270 |
| Intermediate frailty risk | 49,695 (47.9%) |  | 1,824 (58.4%) | 33,354 (53.8%) | 16,341 (39.3%) |  | 912 (57.4%) | 912 (59.4%) |  |
| Charlson Comorbidity Index (continuous) | 5.1 (5.2) |  | 6.3 (5.6) | 5.1 (5.2) | 5.1 (5.1) | 0.195 | 5.6 (5.1) | 6.9 (6.0) | <0.001 |

Note: The ERICCA trial did not have data for index of multiple deprivation, acute coronary syndrome, and hospital frailty risk scores.

Baseline sociodemographic and clinical characteristics of the emulated trial by seven classes are in **Table S7**. The distribution (proportions) of the seven classes were different between the emulated trial and the whole study population, with substantial increase of sample size in Classes 1 (43% versus 25%) and 6 (26% versus 9%) in the emulated trial population, while the other five classes (2-5 and 7) commensurately reduced in sample size, due to the participant profiles of ERICCA trial (old people with CKD) and they were the first two characteristics in the stepwise segmentation by LCA (**Figure 3**). The crude event rates for the composite primary outcome and secondary outcomes between CABG and PCI in the emulated trial by seven classes are in **Table S8**, and the results of IVA in **Table 3** (**lower panel**).

IV was valid for the emulated trial overall and Classes 1, 4, 5, 6, less ideal for Classes 3 and 7 (statistics<10, although significant), not valid for Class 2 (**Table S4**), mainly due to small sample sizes in these three classes. In addition to low event rates, ATEs were unstable or inestimable for any outcome in Class 2 (**Table 3**).

For the primary outcome, the ATE favoured CABG overall 0.55 (0.46 to 0.65) and in Classes 1, 3, and 4, but no difference in Classes 5, 6, and 7. The colour-coded results in **Table 3** indicated heterogeneity in benefits and harms between CABG and PCI in different secondary outcomes (**lower panel,** by column) in Classes 3, 4, 5, and 6 (by row). Patients in Class 1 had consistent results favouring CABG across all outcomes, while all results for Class 7 were null.

When comparing the results between the whole study population (HES, stage 2) and the emulated trial (stage 3), although some point estimates were similar, the emulated trial population tended to have wider 95% CIs than the whole study population due to smaller sample sizes. The composite primary outcome and two secondary outcomes (cardiovascular rehospitalisation and MI) favoured CABG in both populations. CABG consistently led to better results in all outcomes for patients in Class 1 (age>75 years without CKD, with or without the other five high-risk characteristics) in both HES and emulated populations. The rest of results were less consistent.

### Stage 4: sample size and power considerations for a pragmatic trial using Bayesian borrowing

Bayesian borrowing substantially increased power compared with separate analyses, from 67% to 91% (**Figure S5**), and controlled the family-wise error rate (FWER) without adjustment. Under the global alternative hypothesis, the weighted average power for sample sizes of 1500, 2000, 2500 and 3000 was 73%, 82%, 88%, 91%, respectively (**Figure S5**, dashed line and **Table S9**). Therefore, 3000 participants are required for a two-arm RCT with seven classes to achieve 90% power, compared with 990 for a single all-comers trial with a pooled analysis (all clusters combined), 6760 for 7 separate trials recruiting separately until reaching 90% power, and >15000 for one trial recruiting until the average power for class-specific analysis reaches 90%. 2000 participants are required for 80% average power.

In sensitivity analyses with partial alternatives (**Table S10**), average power was robust (still >80%) to 1-4 small clusters having no treatment effect (TE), fairly robust to one large cluster being the only cluster with no TE (74%) or being the only cluster with target TE (72%), but highly sensitive to even the three largest classes having no TE (22%).

## Discussion

### Key findings

This study demonstrated heterogeneous treatment effects (HTE) between CABG and PCI in people with high-risk characteristics requiring multivessel myocardial revascularisation. Latent class analyses segmented the target population into seven non-overlapping classes, each with distinct patterns of high-risk characteristics, different rates of disease progression, and different interactions with revascularisation procedures. Neither the whole study population nor the emulated trial population resolved uncertainty with respect to treatment choices for people with high-risk characteristics requiring multivessel revascularisation. This leads us to suggest that an RCT is warranted to provide high-quality evidence to support individual treatment choices in the target population. Co-production with multiple stakeholders concluded that a pragmatic trial in Bayesian approach with 3000 participants and seven strata would address the remaining uncertainty.

### Clinical relevance

The HTE for the primary and secondary outcomes among classes suggested that an all-comers trial, without stratification, is unlikely to detect potential harms from CABG versus PCI. The heterogeneity of benefits and harms within individual patient classes is an important consideration for patients. Specifically, avoidance of stroke is a primary consideration for many people deciding whether to undergo CABG or PCI for multivessel coronary artery disease, regardless of important benefits including reductions in death and cardiovascular hospitalisation, as shown in Class 3. Shared decision making is central to personalised, values-based treatment decisions, but this is underutilised in myocardial revascularisation. There is an unmet need for data-driven decision support tools that allow patients to match treatment choices to their individual priorities.

All of these issues can be addressed by an RCT of CABG versus PCI in people with high-risk characteristics. Simulation suggested that a trial with 3000 participants for seven strata (classes), allowing dynamic Bayesian borrowing, should be sufficient to address residual uncertainty. This preserves FWER, whereas a stratified frequentist design would not, and enables adequate power even in smaller strata. This sample size is more efficient than seven separate trials or a single trial with 90% power in each individual stratum without borrowing. This design allows co-enrolment with other similar trials, for example, the STICH-3C trial.^25^ The target population is 56.8% of the total service population (wide eligibility) suggests that such a trial would be feasible. It also offers the possibility of using the future trial data to develop a personalised decision support tool for clinicians and patients.

### Strengths and limitations

We used a novel stepwise approach involving well-developed advanced statistical methods, such as applying the target trial emulation framework in longitudinal linked electronic health records, unsupervised machine learning (LCA), causal inference (IVA), and Bayesian borrowing in clinical trial design, to address a knowledge gap of unwarranted variation in treatment preferences and outcomes of multivessel myocardial revascularisation in England. We published the study design, specifying the study rationale and statistical analysis plan, and followed relevant guidelines to conduct and report this study.

LCA was used to identify non-overlapping patient clusters with distinct clinical phenotypes and disease trajectories by considering all the characteristics holistically. Subgroup analysis, commonly used in RCT, is difficult to tease out treatment effects among the intercorrelated clinical characteristics. Compared with other clustering methods, LCA has the advantage of providing posterior probabilities for each patient in each class, to determine class memberships based on the largest probabilities. As an unsupervised learning method, we could not know the true number of classes. To mitigate this limitation, we explored solutions ranged from two to nine latent classes, and set three prespecified criteria to decide the optimal number of classes. Three or four classes as strata in RCT may be more ideal for trial implementation. However, when comparing with eight solutions from LCA, seven classes were considered the optimal solution to represent clinical phenotypes, with small sample sizes in some classes. Our proposed stratified trial design with Bayesian borrowing may overcome these two limitations.

IVA was used to address measured and unmeasured confounders in the TTE. Nonetheless, IVA cannot adjust for all unknown confounders, specifically important determinants of treatment such as the SYNTAX score^3^ and other target vessel characteristics. Therefore, residual confounding is likely. The best way to address unmeasured confounding is through an RCT.

Performing IVA in an emulated trial population using IPD from a high-quality RCT of multivessel revascularisation in 18 UK cardiac centres provides additional information on trial feasibility. Due to a smaller sample size, inestimable and unstable results, and wide confidence intervals were the limitations of estimating the treatment effect in the emulated trial population.

Death is a competing risk for rehospitalisation events. We wanted to use the Fine-Gray model to estimate the sub-distribution hazards. However, when combining competing risk with bootstrapping, we could not get the estimates due to computing capacity even if using HPC. We used Cox regression instead for cause-specific hazards.

The high-risk REVASC study fostered interdisciplinary and international collaborations among health data scientists, trial methodologists, clinicians, and patient and public representatives in complex trial designs for people with complex high-risk characteristics. This study represents a research framework for co-production in trials, transferrable to other disease areas, not just in cardiovascular interventions.

## Conclusion

We conducted this study to investigate the treatment effect of CABG versus PCI in people with high-risk characteristics requiring myocardial revascularisation through target trial emulation using observational data. CABG had favourable results in the composite primary outcome and two secondary outcomes (CV rehospitalisation and MI) in the overall HES population and the emulated trial population. Patients aged >75 years without CKD (Class 1) who received CABG had better results across all clinical outcomes. For the other six patient clusters, heterogeneity in benefits and harms between CABG and PCI existed in different clinical outcomes. These findings will be tested in the future pragmatic stratified RCT in planning.

## Contributions from authors

GJM conceptualised the study. W Liao conducted statistical analyses for stages 1 to 3. IRW, RMT, and W Li conducted the simulation study to determine the sample size and power for the pragmatic trial (stage 4). W Liao and GJM drafted the manuscript, with input from IRW and RMT on stage 4. All authors actively involved in the whole research process, discussed the preliminary findings periodically, reviewed and revised the previous draft versions, and approved the final manuscript for submission.

## Supporting information

High-Risk REVASC. Online supplementary materials

## Data Availability

This study used anonymised NHS patient data. Anyone who wants access to anonymised NHS patient data should directly apply from the data custodian.

## Notes

1. The study design and statistical analysis plan for this main paper have been published in American Heart Journal, 2026 Jun:296:107368. doi: 10.1016/j.ahj.2026.107368.

2. Generative AI was NOT used in writing this article, or any submitted materials for peer review.

## References

1. Lawton JS, Tamis-Holland JE, Bangalore S, et al. 2021 ACC/AHA/SCAI Guideline for Coronary Artery Revascularization: Executive Summary: A Report of the American College of Cardiology/American Heart Association Joint Committee on Clinical Practice Guidelines. Circulation 2022; 145(3): e4–e17.

2. Rao SV, O’Donoghue ML, Ruel M, et al. 2025 ACC/AHA/ACEP/NAEMSP/SCAI Guideline for the Management of Patients With Acute Coronary Syndromes: A Report of the American College of Cardiology/American Heart Association Joint Committee on Clinical Practice Guidelines. Circulation 2025; 151(13): e771–e862.

3. Takahashi K, Serruys PW, Fuster V, et al. Redevelopment and validation of the SYNTAX score II to individualise decision making between percutaneous and surgical revascularisation in patients with complex coronary artery disease: secondary analysis of the multicentre randomised controlled SYNTAXES trial with external cohort validation. Lancet 2020; 396(10260): 1399–412.

4. Head SJ, Milojevic M, Daemen J, et al. Mortality after coronary artery bypass grafting versus percutaneous coronary intervention with stenting for coronary artery disease: a pooled analysis of individual patient data. Lancet 2018; 391(10124): 939–48.

5. Sabatine MS, Bergmark BA, Murphy SA, et al. Percutaneous coronary intervention with drug-eluting stents versus coronary artery bypass grafting in left main coronary artery disease: an individual patient data meta-analysis. Lancet 2021; 398(10318): 2247–57.

6. Rashid M, Lai F, Pathak S, et al. Five-year outcomes of percutaneous coronary intervention and coronary artery bypass grafting for multivessel disease: a national population-based study of regional practice. Eur Heart J Open 2026; 6(2): oeag043.

7. Zheng W, Huang X, Wang X, et al. Impact of multimorbidity patterns on outcomes and treatment in patients with coronary artery disease. Eur Heart J Open 2024; 4(2): oeae009.

8. Ono M, Serruys PW, Hara H, et al. 10-Year Follow-Up After Revascularization in Elderly Patients With Complex Coronary Artery Disease. J Am Coll Cardiol 2021; 77(22): 2761–73.

9. Steigen T, Holm NR, Myrmel T, et al. Age-Stratified Outcome in Treatment of Left Main Coronary Artery Stenosis: A NOBLE Trial Substudy. Cardiology 2021; 146(4): 409–18.

10. Cashin AG, Hansford HJ, Hernan MA, et al. Transparent Reporting of Observational Studies Emulating a Target Trial-The TARGET Statement. JAMA 2025.

11. Liao W, Rashid M, Brookes CL, et al. Study design for an emulated trial of a 2 arm, parallel, stratified, adaptive, RCT of CABG versus PCI in people requiring myocardial revascularization at high risk (High-Risk REVASC). Am Heart J 2026; 296: 107368.

12. Weller BE, Bowen NK, Faubert SJ. Latent Class Analysis: A Guide to Best Practice. Journal of Black Psychology 2020; 46(4): 287–311.

13. Thorndike RL. Who Belongs in the Family? Psychometrika 1953; 18(4): 267–76.

14. Pathak S, Lai FY, Miksza J, et al. Surgical or percutaneous coronary revascularization for heart failure: an in silico model using routinely collected health data to emulate a clinical trial. Eur Heart J 2023; 44(5): 351–64.

15. Roman M, Miksza J, Lai FY, et al. Revascularization in frail patients with acute coronary syndromes: a retrospective longitudinal study. Eur Heart J 2025; 46(6): 535–47.

16. Baig SS, Altman DG, Taggart DP. Major geographical variations in elective coronary revascularization by stents or surgery in England. Eur J Cardiothorac Surg 2015; 47(5): 855–9.

17. Walker V, Sanderson E, Levin MG, Damraurer SM, Feeney T, Davies NM. Reading and conducting instrumental variable studies: guide, glossary, and checklist. BMJ 2024; 387: e078093.

18. Fu R, Kim SJ. Inferring causality from observational studies: the role of instrumental variable analysis. Kidney Int 2021; 99(6): 1303–8.

19. Terza JV, Basu A, Rathouz PJ. Two-stage residual inclusion estimation: addressing endogeneity in health econometric modeling. J Health Econ 2008; 27(3): 531–43.

20. Bidulka P, Lugo-Palacios DG, Carroll O, et al. Comparative effectiveness of second line oral antidiabetic treatments among people with type 2 diabetes mellitus: emulation of a target trial using routinely collected health data. BMJ 2024; 385: e077097.

21. Martinez-Camblor P, Mackenzie T, Staiger DO, Goodney PP, O’Malley AJ. Adjusting for bias introduced by instrumental variable estimation in the Cox proportional hazards model. Biostatistics 2019; 20(1): 80–96.

22. Hausenloy DJ, Candilio L, Evans R, et al. Remote Ischemic Preconditioning and Outcomes of Cardiac Surgery. N Engl J Med 2015; 373(15): 1408–17.

23. Ouma LO, Grayling MJ, Wason JMS, Zheng H. Bayesian modelling strategies for borrowing of information in randomised basket trials. J R Stat Soc Ser C Appl Stat 2022; 71(5): 2014–37.

24. Turner RM, Turkova A, Moore CL, et al. Borrowing information across patient subgroups in clinical trials, with application to a paediatric trial. BMC Med Res Methodol 2022; 22(1): 49.

25. Fremes SE, Marquis-Gravel G, Gaudino MFL, et al. STICH3C: Rationale and Study Protocol. Circ Cardiovasc Interv 2023; 16(8): e012527.

