## Supplementary material for "Target trial emulation for a two-arm, parallel, stratified pragmatic RCT of CABG versus PCI in people with high-risk characteristics requiring myocardial revascularisation (High-Risk REVASC)": High-Risk REVASC. Online supplementary materials

Corresponding Author

Dr Weiqi Liao,.

Division of Cardiovascular Sciences, School of Medical Sciences, University of Leicester

Clinical Sciences Wing, Glenfield Hospital, Leicester, LE3 9QP

medRxiv

medRxiv

### Supplementary materials – Methods

#### Sample size estimation and power consideration for the target pragmatic trial

##### Parameters and assumptions about the data generating model (DGM)

The proportions (prevalence) for classes 1-7 in the whole study population (HES) were 0.25, 0.04, 0.06, 0.27, 0.24, 0.09, and 0.05, respectively. The 5-year cumulative incidence of the composite primary outcome (all-cause mortality or cardiovascular rehospitalisation) in the control group (PCI) in the whole study population (HES, stage 2) for classes 1-7 was 0.623, 0.490, 0.398, 0.609, 0.392, 0.720, and 0.689, respectively. Weibull regression was used as the outcome model, with the same shape parameter (0.48) derived from the 5-year incidence of the composite outcome for both arms and across classes. The primary outcome for the target trial (same as above) was estimated assuming a minimum of 4 years of follow-up. The target treatment effect (TE) was set as 0.7. The expected cross-over from previous studies was 8%. The target TE with cross-overs was 0.739 ($e^{\log(0.7)\times(1-0.08)^2}$).

The R package *bayesmeta* was used to estimate the required sample size for the planned Bayesian borrowing analysis. The model assumed TEs were random across classes. Heterogeneity of TEs between classes (τ) had a half-normal prior with scale parameter 0.26, i.e. $\tau \sim N(0,0.26^2)I[0,\infty]$, which was an empirically derived distribution (https://arxiv.org/abs/2306.11468), median $\tau = 0.175$ and $P(\tau > 0.26) = 0.32$. The posterior probabilities of each class-specific TE being <0 were extracted. If the posterior probability was >0.975 (2.5% one-sided error rate, or $1-\alpha/2$ in general), then it was interpreted as a significant result in a class. The target power was 90%.

##### Computational approach

The overall idea was to take various candidate sample sizes N, use simulation to estimate the power, and refine the sample size if necessary until the required power is achieved.

We started with an initial guess for the sample size N between the “class-specific” and “overall” sample sizes. Then we used N and the DGM (described above) to generate individual participant

medRxiv

data for the seven classes. A Cox model was fitted separately for each class, extracting the log HR and its SE. Simulated data were analysed either by separate analyses or by a Bayesian hierarchical model. This simulation process was repeated 1000 times for each set of true TEs (eight sets, below). The global null (1 below) simulations were used to identify the class-specific P-value threshold that controls the family-wise type 1 error rate (FWER) at the 5% level, and this P-value threshold was used to evaluate TE in each class. Performance measures for each set of true TEs were computed. If the average power differed from the target power, we updated N and restarted the simulation process.

Eight sets of true subgroup-specific TEs:

1. Global null: All TEs are null (i.e. log HR = 0, HR = 1)
2. Global alternative: all TEs are the target TE
3. Partial alternative: some nulls and some non-nulls (non-global nulls: treatment works in some subgroups). The non-null value will equal the target.
    a. one null (the smallest class) + six non-nulls
    b. one null (the largest class) + six non-nulls
    c. six nulls + one non-null (the smallest class)
    d. six nulls + one non-null (the largest class)
    e. three nulls (the 3 largest) + four non-nulls
    f. four nulls + three non-nulls (the 3 largest)

medRxiv

### Performance measures

The overall performance measures were described in the main paper. Class-specific performance measures included (1) type 1 error rates per class in which TE is null and (2) power per class in which TE is non-null.

### Supplementary materials – Tables

Table S1 – Model summary statistics from two to nine classes using latent class analysis (stage 1)

| Number of classes | Number of estimated parameters | Residual degrees of freedom | Maximum log-likelihood | AIC | BIC | $G^2$ | $\chi^2$ | Entropy |
|---|---|---|---|---|---|---|---|---|
| 2 | 15 | 112 | -358882.5 | 717794.9 | 717938.2 | 24482.7 | 20136.0 | 3.461861 |
| 3 | 23 | 104 | -355768.3 | 711582.7 | 711802.3 | 18254.5 | 15012.6 | 3.431213 |
| 4 | 31 | 96 | -353724 | 707509.9 | 707805.9 | 14165.7 | 12621.1 | 3.411579 |
| 5 | 39 | 88 | -351112.5 | 702303 | 702675.4 | 8942.8 | 8356.8 | 3.386624 |
| 6 | 47 | 80 | -349785.8 | 699665.6 | 700114.4 | 6289.4 | 5491.5 | 3.374153 |
| 7 | 55 | 72 | -347145.2 | 694400.4 | 694925.6 | 1008.2 | 1062.2 | 3.348566 |
| 8 | 63 | 64 | -346845 | 693816 | 694417.6 | 407.8 | 421.5 | 3.345865 |
| 9 | 71 | 56 | -346755.5 | 693653 | 694331 | 228.8 | 236.3 | 3.344937 |

Note: AIC (Akaike Information Criterion), BIC (Bayesian Information Criterion), $G^2$ (Likelihood ratio/deviance statistic), $\chi^2$ (Chi-square goodness of fit)

medRxiv

Table S2 – Frequencies and proportions of predicted class membership for each class in solutions with two to nine classes using latent class analysis (stage 1)

| Number of latent classes | Class 1 | Class 2 | Class 3 | Class 4 | Class 5 | Class 6 | Class 7 | Class 8 | Class 9 |
|---|---|---|---|---|---|---|---|---|---|
| 2 | 70,990 (68.5%) | 32,680 (31.5%) | | | | | | | |
| 3 | 29,228 (28.2%) | 59,076 (57.0%) | 15,366 (14.8%) | | | | | | |
| 4 | 19,863 (19.2%) | 40,990 (39.5%) | 19,626 (18.9%) | 23,191 (22.4%) | | | | | |
| 5 | 11,786 (11.4%) | 8,492 (8.2%) | 21,257 (20.5%) | 32,191 (31.1%) | 29,944 (28.9%) | | | | |
| 6 | 30,343 (29.3%) | 21,000 (20.3%) | 11,563 (11.2%) | 25,950 (25.0%) | 8,947 (8.6%) | 5,867 (5.7%) | | | |
| 7 | 25,950 (25.0%) | 4,040 (3.9%) | 6,477 (6.2%) | 28,325 (27.3%) | 24,698 (23.8%) | 8,947 (8.6%) | 5,233 (5.0%) | | |
| 8 | 5,361 (5.2%) | 22,224 (21.4%) | 11,172 (10.8%) | 28,325 (27.3%) | 3,453 (3.3%) | 6,010 (5.8%) | 20,590 (19.9%) | 6,535 (6.3%) | |
| 9 | 4,175 (4.0%) | 8,742 (8.4%) | 14,046 (13.5%) | 3,244 (3.1%) | 3,453 (3.3%) | 35,311 (34.1%) | 8,472 (8.2%) | 5,637 (5.4%) | 20,590 (19.9%) |

Note: n (%) in the table above, N = 103,670 eligible patients from hospital episode statistics (HES).

medRxiv

Table S3 – Crude event rates for the composite primary outcome and secondary outcomes at 5 years in the whole study population (HES) by seven latent classes and two revascularisation approaches

| HES | | N | All-cause mortality or cardiovascular hospitalisation | All-cause mortality | Cardiovascular mortality | Cardiovascular hospitalisation | Myocardial infarction | Heart failure hospitalisation | Stroke |
|---|---|---|---|---|---|---|---|---|---|
| Class 1 | CABG | 16,981 | 8,044 (47.4%) | 4,454 (26.2%) | 3,486 (20.5%) | 5,223 (30.8%) | 480 (2.8%) | 1,053 (6.2%) | 484 (2.9%) |
| | PCI | 8,969 | 5,586 (62.3%) | 3,059 (34.1%) | 2,459 (27.4%) | 3,819 (42.6%) | 673 (7.5%) | 623 (6.9%) | 288 (3.2%) |
| Class 2 | CABG | 3,536 | 1,196 (33.8%) | 459 (13.0%) | 406 (11.5%) | 853 (24.1%) | 52 (1.5%) | 139 (3.9%) | 27 (0.8%) |
| | PCI | 504 | 247 (49.0%) | 67 (13.3%) | 58 (11.5%) | 207 (41.1%) | 23 (4.6%) | 30 (6.0%) | 7 (1.4%) |
| Class 3 | CABG | 4,339 | 1,082 (24.9%) | 231 (5.3%) | 186 (4.3%) | 904 (20.8%) | 80 (1.8%) | 55 (1.3%) | 52 (1.2%) |
| | PCI | 2,138 | 850 (39.8%) | 114 (5.3%) | 88 (4.1%) | 778 (36.4%) | 82 (3.8%) | 14 (0.7%) | 13 (0.6%) |
| Class 4 | CABG | 18,294 | 7,999 (43.7%) | 2,362 (12.9%) | 1,745 (9.5%) | 6,672 (36.5%) | 694 (3.8%) | 1,039 (5.7%) | 642 (3.5%) |
| | PCI | 10,031 | 6,104 (60.9%) | 1,424 (14.2%) | 1,041 (10.4%) | 5,439 (54.2%) | 1,035 (10.3%) | 549 (5.5%) | 341 (3.4%) |
| Class 5 | CABG | 7,855 | 1,832 (23.3%) | 478 (6.1%) | 393 (5.0%) | 1,435 (18.3%) | 214 (2.7%) | 79 (1.0%) | 51 (0.6%) |
| | PCI | 16,843 | 6,598 (39.2%) | 1,183 (7.0%) | 960 (5.7%) | 5,759 (34.2%) | 1,063 (6.3%) | 219 (1.3%) | 131 (0.8%) |
| Class 6 | CABG | 6,805 | 3,763 (55.3%) | 2,388 (35.1%) | 2,078 (30.5%) | 2,177 (32.0%) | 229 (3.4%) | 572 (8.4%) | 177 (2.6%) |
| | PCI | 2,142 | 1,542 (72.0%) | 987 (46.1%) | 869 (40.6%) | 1,094 (51.1%) | 236 (11.0%) | 260 (12.1%) | 65 (3.0%) |
| Class 7 | CABG | 4,238 | 2,087 (49.2%) | 717 (16.9%) | 557 (13.1%) | 1,705 (40.2%) | 145 (3.4%) | 198 (4.7%) | 152 (3.6%) |
| | PCI | 995 | 686 (68.9%) | 241 (24.2%) | 200 (20.1%) | 556 (55.9%) | 98 (9.8%) | 52 (5.2%) | 41 (4.1%) |
| All | CABG | 62,048 | 26,003 (41.9%) | 11,089 (17.9%) | 8,851 (14.3%) | 18,969 (30.6%) | 1,894 (3.1%) | 3,135 (5.1%) | 1,585 (2.6%) |
| | PCI | 41,622 | 21,613 (51.9%) | 7,075 (17.0%) | 5,675 (13.6%) | 17,652 (42.4%) | 3,210 (7.7%) | 1,747 (4.2%) | 886 (2.1%) |

Table S4 – Strength of the instrumental variable (regional variation in standardised CABG rates) in the whole study population (HES, stage 2) and the emulated trial (ERICCA matched to HES, stage 3)

| | HES | P value (HES) | Emulated trial | P value (emulated) |
|---|---|---|---|---|
| **All** | 3502.155 | <0.001 | 156.517 | <0.001 |
| **Class 1** | 1188.095 | <0.001 | 91.678 | <0.001 |
| **Class 2** | 40.011 | <0.001 | 0.001 | 0.971 |
| **Class 3** | 206.665 | <0.001 | 8.558 | 0.003 |
| **Class 4** | 805.238 | <0.001 | 14.431 | <0.001 |
| **Class 5** | 733.686 | <0.001 | 25.946 | <0.001 |
| **Class 6** | 288.187 | <0.001 | 24.070 | <0.001 |
| **Class 7** | 187.636 | <0.001 | 7.548 | 0.006 |

Note: HES (hospital episode statistics), ERICCA (doi: 10.1056/NEJMoa1413534)

medRxiv

Table S5 – Variables used for propensity score matching (nearest neighbour) between the ERICCA trial and the HES CABG arm for the emulated trial (stage 3)

| Variables | | ERICCA | HES (CABG) | % bias | % reduct \|bias\| | t | P value |
|---|---|---|---|---|---|---|---|
| **Age** | Unmatched (U) | 76.2 | 68.8 | 88 | | 29.58 | <0.001 |
| | Matched (M) | 76.2 | 76.2 | -0.1 | 99.9 | -0.02 | 0.985 |
| **Female sex** | U | 28.4% | 25.9% | 5.7 | | 2.29 | 0.022 |
| | M | 28.4% | 29.8% | -3.1 | 45.7 | -0.86 | 0.39 |
| **Heart failure** | U | 30.0% | 22.1% | 17.9 | | 7.4 | <0.001 |
| | M | 29.8% | 30.9% | -2.3 | 87.1 | -0.62 | 0.537 |
| **Hypertension** | U | 75.3% | 76.0% | -1.7 | | -0.66 | 0.51 |
| | M | 75.4% | 75.9% | -1.2 | 29.4 | -0.33 | 0.741 |
| **Diabetes** | U | 25.9% | 27.4% | -3.4 | | -1.33 | 0.184 |
| | M | 26.0% | 24.9% | 2.6 | 24.6 | 0.73 | 0.463 |
| **Chronic kidney disease** | U | 28.5% | 11.0% | 45.2 | | 21.85 | <0.001 |
| | M | 28.3% | 27.9% | 1 | 97.8 | 0.24 | 0.813 |
| **Cerebrovascular disease** | U | 11.5% | 5.8% | 20.4 | | 9.51 | <0.001 |
| | M | 11.3% | 11.1% | 0.4 | 97.8 | 0.11 | 0.91 |
| **Myocardial infarction** | U | 39.9% | 35.5% | 9.1 | | 3.62 | <0.001 |
| | M | 39.9% | 38.7% | 2.3 | 74.2 | 0.65 | 0.513 |

| | | | | | | | |
|---|---|---|---|---|---|---|---|
| **Peripheral vascular disease** | U | 7.6% | 10.3% | -9.6 | | -3.54 | <0.001 |
| | M | 7.6% | 6.6% | 3.5 | 63 | 1.1 | 0.27 |
| **Lipidaemia** | U | 70.4% | 67.3% | 6.7 | | 2.6 | 0.009 |
| | M | 70.3% | 71.0% | -1.5 | 77.6 | -0.43 | 0.668 |
| **White** | U | 94.4% | 91.1% | 12.5 | | 4.52 | <0.001 |
| | M | 94.3% | 95.3% | -3.6 | 70.9 | -1.2 | 0.231 |
| **Asian** | U | 4.8% | 6.4% | -6.8 | | -2.52 | 0.012 |
| | M | 4.8% | 3.8% | 4.4 | 35.3 | 1.39 | 0.164 |
| **Black** | U | 0.5% | 0.7% | -3 | | -1.08 | 0.281 |
| | M | 0.5% | 0.4% | 0.8 | 72.9 | 0.26 | 0.796 |

Note: "Unmatched (U)" row means the results before propensity score matching; "Matched (M)" row means the results after propensity score matching.

medRxiv

Table S6 – Variables used for propensity score matching (nearest neighbour) between the ERICCA trial and the HES PCI arm for the emulated trial (stage 3)

| Variables | | ERICCA | HES (PCI) | % bias | % reduct \|bias\| | t | P value |
|---|---|---|---|---|---|---|---|
| **Age** | Unmatched (U) | 76.2 | 65.5 | 113 | | 36.31 | <0.001 |
| | Matched (M) | 76.1 | 76.7 | -6.4 | 94.3 | -2.26 | 0.024 |
| **Female sex** | U | 28.4% | 26.9% | 3.4 | | 1.35 | 0.177 |
| | M | 28.6% | 27.5% | 2.5 | 27.7 | 0.68 | 0.495 |
| **Heart failure** | U | 30.0% | 8.2% | 57.6 | | 30.07 | <0.001 |
| | M | 28.5% | 27.7% | 2.2 | 96.1 | 0.52 | 0.602 |
| **Hypertension** | U | 75.3% | 42.1% | 71.5 | | 26.45 | <0.001 |
| | M | 74.9% | 77.1% | -4.9 | 93.1 | -1.48 | 0.139 |
| **Diabetes** | U | 25.9% | 16.3% | 23.8 | | 10.17 | <0.001 |
| | M | 25.9% | 25.5% | 1 | 96 | 0.25 | 0.804 |
| **Chronic kidney disease** | U | 28.5% | 5.1% | 65.7 | | 39.22 | <0.001 |
| | M | 26.8% | 24.5% | 6.2 | 90.5 | 1.4 | 0.16 |
| **Cerebrovascular disease** | U | 11.5% | 1.3% | 42.6 | | 31.41 | <0.001 |
| | M | 9.2% | 7.0% | 9.2 | 78.3 | 2.24 | 0.025 |
| **Myocardial infarction** | U | 39.9% | 34.3% | 11.6 | | 4.6 | <0.001 |
| | M | 40.2% | 42.6% | -5 | 56.8 | -1.36 | 0.175 |

| | | | | | | | |
|---|---|---|---|---|---|---|---|
| **Peripheral vascular disease** | U | 7.6% | 4.0% | 15.6 | | 7.2 | <0.001 |
| | M | 7.7% | 7.2% | 2.2 | 85.7 | 0.55 | 0.584 |
| **Lipidaemia** | U | 70.4% | 29.8% | 88.9 | | 34.81 | <0.001 |
| | M | 69.5% | 69.8% | -0.6 | 99.4 | -0.16 | 0.875 |
| **White** | U | 94.4% | 90.4% | 14.9 | | 5.29 | <0.001 |
| | M | 94.2% | 94.6% | -1.5 | 90.1 | -0.47 | 0.638 |
| **Asian** | U | 4.8% | 6.7% | -8.2 | | -3.02 | 0.003 |
| | M | 4.9% | 4.9% | 0.3 | 96.6 | 0.08 | 0.934 |
| **Black** | U | 0.5% | 0.9% | -5.1 | | -1.77 | 0.076 |
| | M | 0.5% | 0.3% | 3.1 | 39.6 | 1.16 | 0.247 |

Note: "Unmatched (U)" row means the results before propensity score matching; "Matched (M)" row means the results after propensity score matching.

medRxiv

Table S7 – Baseline sociodemographic and clinical characteristics of the emulated trial by seven latent classes (stage 3)

| | Class 1 | Class 2 | Class 3 | Class 4 | Class 5 | Class 6 | Class 7 | Total | P value |
|---|---|---|---|---|---|---|---|---|---|
| N (%) | 1,361 (43.6%) | 69 (2.2%) | 92 (2.9%) | 467 (14.9%) | 269 (8.6%) | 820 (26.2%) | 46 (1.5%) | 3,124 (100.0%) | |
| Revascularisation approach | | | | | | | | | |
| PCI | 690 (50.7%) | 17 (24.6%) | 33 (35.9%) | 229 (49.0%) | 169 (62.8%) | 377 (46.0%) | 21 (45.7%) | 1,536 (49.2%) | <0.001 |
| CABG | 671 (49.3%) | 52 (75.4%) | 59 (64.1%) | 238 (51.0%) | 100 (37.2%) | 443 (54.0%) | 25 (54.3%) | 1,588 (50.8%) | |
| Age | 81.2 (4.1) | 67.9 (5.2) | 69.7 (4.7) | 69.1 (5.1) | 67.0 (6.7) | 77.7 (6.8) | 68.4 (5.7) | 76.4 (7.7) | <0.001 |
| ≤75 years | 0 (0.0%) | 69 (100.0%) | 92 (100.0%) | 467 (100.0%) | 269 (100.0%) | 253 (30.9%) | 46 (100.0%) | 1,196 (38.3%) | <0.001 |
| >75 years | 1,361 (100.0%) | 0 (0.0%) | 0 (0.0%) | 0 (0.0%) | 0 (0.0%) | 567 (69.1%) | 0 (0.0%) | 1,928 (61.7%) | |
| Sex | | | | | | | | | |
| Male | 944 (69.4%) | 54 (78.3%) | 0 (0.0%) | 366 (78.4%) | 219 (81.4%) | 612 (74.6%) | 33 (71.7%) | 2,228 (71.3%) | <0.001 |
| Female | 417 (30.6%) | 15 (21.7%) | 92 (100.0%) | 101 (21.6%) | 50 (18.6%) | 208 (25.4%) | 13 (28.3%) | 896 (28.7%) | |
| Ethnicity | | | | | | | | | |
| White | 1,188 (96.4%) | 61 (93.8%) | 77 (98.7%) | 407 (93.6%) | 229 (93.5%) | 696 (91.8%) | 41 (95.3%) | 2,699 (94.5%) | <0.001 |
| Asian | 40 (3.2%) | 1 (1.5%) | 1 (1.3%) | 24 (5.5%) | 15 (6.1%) | 54 (7.1%) | 1 (2.3%) | 136 (4.8%) | |
| Black | 1 (0.1%) | 2 (3.1%) | 0 (0.0%) | 3 (0.7%) | 1 (0.4%) | 3 (0.4%) | 1 (2.3%) | 11 (0.4%) | |
| Mixed/others | 4 (0.3%) | 1 (1.5%) | 0 (0.0%) | 1 (0.2%) | 0 (0.0%) | 5 (0.7%) | 0 (0.0%) | 11 (0.4%) | |
| Index of Multiple Deprivation score (continuous) | 17.5 (13.6) | 19.7 (14.2) | 19.0 (15.0) | 21.7 (15.6) | 20.6 (15.0) | 20.2 (14.6) | 24.8 (16.1) | 19.3 (14.5) | <0.001 |
| Index of Multiple Deprivation (quintile) | | | | | | | | | |
| Least deprived quintile (0-20%) | 375 (27.6%) | 15 (21.7%) | 15 (16.3%) | 94 (20.1%) | 59 (21.9%) | 170 (20.7%) | 7 (15.2%) | 735 (23.5%) | <0.001 |
| Less deprived quintile | 307 (22.6%) | 15 (21.7%) | 26 (28.3%) | 84 (18.0%) | 48 (17.8%) | 160 (19.5%) | 6 (13.0%) | 646 (20.7%) | |
| Middle quintile | 222 (16.3%) | 10 (14.5%) | 17 (18.5%) | 91 (19.5%) | 43 (16.0%) | 159 (19.4%) | 8 (17.4%) | 550 (17.6%) | |
| More deprived quintile | 205 (15.1%) | 13 (18.8%) | 6 (6.5%) | 89 (19.1%) | 52 (19.3%) | 168 (20.5%) | 12 (26.1%) | 545 (17.4%) | |
| Most deprived quintile | 214 (15.7%) | 14 (20.3%) | 21 (22.8%) | 105 (22.5%) | 58 (21.6%) | 146 (17.8%) | 13 (28.3%) | 571 (18.3%) | |
| Unknown | 38 (2.8%) | 2 (2.9%) | 7 (7.6%) | 4 (0.9%) | 9 (3.3%) | 17 (2.1%) | 0 (0.0%) | 77 (2.5%) | |
| Hypertension | | | | | | | | | |
| No | 343 (25.2%) | 18 (26.1%) | 32 (34.8%) | 132 (28.3%) | 96 (35.7%) | 107 (13.0%) | 6 (13.0%) | 734 (23.5%) | <0.001 |

medRxiv

| | | | | | | | | | |
|---|---|---|---|---|---|---|---|---|---|
| Yes | 1,018 (74.8%) | 51 (73.9%) | 60 (65.2%) | 335 (71.7%) | 173 (64.3%) | 713 (87.0%) | 40 (87.0%) | 2,390 (76.5%) | |
| Myocardial infarction | | | | | | | | | |
| No | 875 (64.3%) | 43 (62.3%) | 81 (88.0%) | 295 (63.2%) | 102 (37.9%) | 430 (52.4%) | 28 (60.9%) | 1,854 (59.3%) | <0.001 |
| Yes | 486 (35.7%) | 26 (37.7%) | 11 (12.0%) | 172 (36.8%) | 167 (62.1%) | 390 (47.6%) | 18 (39.1%) | 1,270 (40.7%) | |
| Acute Coronary Syndrome | | | | | | | | | |
| No | 932 (68.5%) | 69 (100.0%) | 92 (100.0%) | 330 (70.7%) | 0 (0.0%) | 529 (64.5%) | 33 (71.7%) | 1,985 (63.5%) | <0.001 |
| Yes | 429 (31.5%) | 0 (0.0%) | 0 (0.0%) | 137 (29.3%) | 269 (100.0%) | 291 (35.5%) | 13 (28.3%) | 1,139 (36.5%) | |
| Heart failure | | | | | | | | | |
| No | 1,024 (75.2%) | 0 (0.0%) | 92 (100.0%) | 369 (79.0%) | 230 (85.5%) | 467 (57.0%) | 27 (58.7%) | 2,209 (70.7%) | <0.001 |
| Yes | 337 (24.8%) | 69 (100.0%) | 0 (0.0%) | 98 (21.0%) | 39 (14.5%) | 353 (43.0%) | 19 (41.3%) | 915 (29.3%) | |
| Chronic kidney disease | | | | | | | | | |
| No | 1,361 (100.0%) | 69 (100.0%) | 92 (100.0%) | 467 (100.0%) | 269 (100.0%) | 0 (0.0%) | 46 (100.0%) | 2,304 (73.8%) | <0.001 |
| Yes | 0 (0.0%) | 0 (0.0%) | 0 (0.0%) | 0 (0.0%) | 0 (0.0%) | 820 (100.0%) | 0 (0.0%) | 820 (26.2%) | |
| Peripheral vascular disease | | | | | | | | | |
| No | 1,288 (94.6%) | 69 (100.0%) | 92 (100.0%) | 467 (100.0%) | 269 (100.0%) | 723 (88.2%) | 0 (0.0%) | 2,908 (93.1%) | <0.001 |
| Yes | 73 (5.4%) | 0 (0.0%) | 0 (0.0%) | 0 (0.0%) | 0 (0.0%) | 97 (11.8%) | 46 (100.0%) | 216 (6.9%) | |
| Diabetes | | | | | | | | | |
| No | 1,093 (80.3%) | 53 (76.8%) | 81 (88.0%) | 344 (73.7%) | 235 (87.4%) | 505 (61.6%) | 26 (56.5%) | 2,337 (74.8%) | <0.001 |
| Yes | 268 (19.7%) | 16 (23.2%) | 11 (12.0%) | 123 (26.3%) | 34 (12.6%) | 315 (38.4%) | 20 (43.5%) | 787 (25.2%) | |
| Cerebrovascular disease | | | | | | | | | |
| No | 1,255 (92.2%) | 66 (95.7%) | 88 (95.7%) | 416 (89.1%) | 255 (94.8%) | 727 (88.7%) | 32 (69.6%) | 2,839 (90.9%) | <0.001 |
| Yes | 106 (7.8%) | 3 (4.3%) | 4 (4.3%) | 51 (10.9%) | 14 (5.2%) | 93 (11.3%) | 14 (30.4%) | 285 (9.1%) | |
| Lipidaemia | | | | | | | | | |
| No | 445 (32.7%) | 23 (33.3%) | 15 (16.3%) | 118 (25.3%) | 74 (27.5%) | 242 (29.5%) | 7 (15.2%) | 924 (29.6%) | <0.001 |
| Yes | 916 (67.3%) | 46 (66.7%) | 77 (83.7%) | 349 (74.7%) | 195 (72.5%) | 578 (70.5%) | 39 (84.8%) | 2,200 (70.4%) | |
| Hospital frailty risk scores (continuous) | 6.3 (4.5) | 2.1 (1.7) | 1.3 (1.3) | 9.3 (2.8) | 1.5 (1.6) | 7.6 (4.2) | 6.3 (4.0) | 6.4 (4.5) | <0.001 |
| Frailty risk (categorical) | | | | | | | | | |

medRxiv

| | | | | | | | | | |
|---|---|---|---|---|---|---|---|---|---|
| Low frailty risk | 600 (44.1%) | 69 (100.0%) | 92 (100.0%) | 0 (0.0%) | 269 (100.0%) | 251 (30.6%) | 19 (41.3%) | 1,300 (41.6%) | <0.001 |
| Intermediate frailty risk | 761 (55.9%) | 0 (0.0%) | 0 (0.0%) | 467 (100.0%) | 0 (0.0%) | 569 (69.4%) | 27 (58.7%) | 1,824 (58.4%) | |
| Charlson Comorbidity Index (continuous) | 5.2 (5.0) | 4.4 (3.9) | 2.4 (3.3) | 7.1 (6.0) | 3.5 (3.4) | 8.9 (5.9) | 8.5 (5.6) | 6.3 (5.6) | <0.001 |

medRxiv

Table S8 – Crude event rates for the composite primary outcome and secondary outcomes at 5 years in the emulated trial by seven latent classes and two revascularisation approaches (stage 3)

| Emulated | | N | All-cause mortality or cardiovascular hospitalisation | All-cause mortality | Cardiovascular mortality | Cardiovascular hospitalisation | Myocardial infarction | Heart failure hospitalisation | Stroke |
|---|---|---|---|---|---|---|---|---|---|
| Class 1 | CABG | 671 | 356 (53.1%) | 204 (30.4%) | 158 (23.5%) | 225 (33.5%) | 19 (2.8%) | 52 (7.7%) | 24 (3.6%) |
| | PCI | 690 | 473 (68.6%) | 292 (42.3%) | 248 (35.9%) | 325 (47.1%) | 56 (8.1%) | 65 (9.4%) | 25 (3.6%) |
| Class 2 | CABG | 52 | 15 (28.8%) | 4 (7.7%) | 3 (5.8%) | 14 (26.9%) | 0 (0.0%) | 2 (3.8%) | 0 (0.0%) |
| | PCI | 17 | 9 (52.9%) | 3 (17.6%) | 3 (17.6%) | 6 (35.3%) | 0 (0.0%) | 1 (5.9%) | 1 (5.9%) |
| Class 3 | CABG | 59 | 16 (27.1%) | 3 (5.1%) | 2 (3.4%) | 13 (22.0%) | 2 (3.4%) | 2 (3.4%) | 1 (1.7%) |
| | PCI | 33 | 13 (39.4%) | 2 (6.1%) | 2 (6.1%) | 12 (36.4%) | 1 (3.0%) | 0 (0.0%) | 0 (0.0%) |
| Class 4 | CABG | 238 | 106 (44.5%) | 38 (16.0%) | 25 (10.5%) | 81 (34.0%) | 9 (3.8%) | 6 (2.5%) | 4 (1.7%) |
| | PCI | 229 | 159 (69.4%) | 53 (23.1%) | 36 (15.7%) | 136 (59.4%) | 26 (11.4%) | 16 (7.0%) | 4 (1.7%) |
| Class 5 | CABG | 100 | 24 (24.0%) | 4 (4.0%) | 4 (4.0%) | 21 (21.0%) | 4 (4.0%) | 1 (1.0%) | 0 (0.0%) |
| | PCI | 169 | 64 (37.9%) | 12 (7.1%) | 7 (4.1%) | 55 (32.5%) | 10 (5.9%) | 1 (0.6%) | 1 (0.6%) |
| Class 6 | CABG | 443 | 277 (62.5%) | 193 (43.6%) | 172 (38.8%) | 146 (33.0%) | 16 (3.6%) | 37 (8.4%) | 20 (4.5%) |
| | PCI | 377 | 295 (78.2%) | 201 (53.3%) | 182 (48.3%) | 210 (55.7%) | 51 (13.5%) | 61 (16.2%) | 15 (4.0%) |
| Class 7 | CABG | 25 | 12 (48.0%) | 7 (28.0%) | 7 (28.0%) | 7 (28.0%) | 0 (0.0%) | 1 (4.0%) | 0 (0.0%) |
| | PCI | 21 | 18 (85.7%) | 13 (61.9%) | 12 (57.1%) | 10 (47.6%) | 1 (4.8%) | 2 (9.5%) | 0 (0.0%) |
| All | CABG | 1,588 | 806 (50.8%) | 453 (28.5%) | 371 (23.4%) | 507 (31.9%) | 50 (3.1%) | 101 (6.4%) | 49 (3.1%) |
| | PCI | 1,536 | 1,031 (67.1%) | 576 (37.5%) | 490 (31.9%) | 754 (49.1%) | 145 (9.4%) | 146 (9.5%) | 46 (3.0%) |

Table S9 – Power for each individual stratum (latent class) under different scenarios for the target pragmatic trial (stage 4)

| Data generation model | N | Analysis | Class 1 | Class 2 | Class 3 | Class 4 | Class 5 | Class 6 | Class 7 | FWER | Weighted average power |
|---|---|---|---|---|---|---|---|---|---|---|---|
| Global null | | | | | | | | | | | |
| | 1500 | Separate | 2.6% | 2.3% | 2.5% | 2.6% | 2.2% | 2.5% | 2.3% | 15.6% | NaN |
| | | Borrow | 1.1% | 0.0% | 0.1% | 1.3% | 0.6% | 0.4% | 0.2% | 2.7% | NaN |
| Global alternative | | | | | | | | | | | |
| | 1500 | Separate | 56.2% | 9.8% | 11.5% | 58.6% | 35.8% | 27.0% | 15.9% | 0.0% | 42.8% |
| | | Borrow | 83.1% | 42.3% | 45.4% | 83.2% | 70.3% | 63.8% | 52.2% | 0.0% | 72.9% |
| | 2000 | Separate | 68.0% | 12.3% | 13.8% | 70.9% | 44.8% | 36.4% | 19.3% | 0.0% | 52.5% |
| | | Borrow | 89.9% | 54.2% | 58.0% | 90.9% | 80.2% | 74.9% | 63.7% | 0.0% | 81.8% |
| | 2500 | Separate | 78.0% | 14.6% | 17.1% | 78.4% | 54.8% | 43.2% | 24.5% | 0.0% | 60.5% |
| | | Borrow | 94.5% | 63.2% | 66.3% | 94.8% | 86.8% | 82.0% | 71.7% | 0.0% | 87.5% |
| | 3000 | Separate | 85.0% | 17.1% | 21.3% | 86.9% | 61.7% | 49.0% | 28.8% | 0.0% | 67.3% |
| | | Borrow | 96.8% | 69.8% | 73.4% | 97.0% | 90.8% | 85.9% | 77.3% | 0.0% | 91.0% |

Note: FWER – Family-wise error rate

medRxiv

Table S10 – Power for each individual stratum (latent class) under six scenarios when the total sample size is 3000 (sensitivity analysis, stage 4)

| Data generation model | N | Analysis | Class 1 | Class 2 | Class 3 | Class 4 | Class 5 | Class 6 | Class 7 | Family-wise error rate | Weighted average power |
|---|---|---|---|---|---|---|---|---|---|---|---|
| Partial alternative 1: only latent class with the ***smallest*** sample size (2) is null | | | | | | | | | | | |
| | 3000 | Separate | 84.9% | 2.0% | 20.6% | 86.5% | 62.9% | 49.3% | 29.9% | 2.0% | 69.6% |
| | | Borrow | 96.0% | 33.7% | 64.9% | 96.4% | 89.1% | 84.0% | 72.1% | 33.7% | 90.1% |
| Partial alternative 2: only latent class with the ***largest*** sample size (4) is null | | | | | | | | | | | |
| | 3000 | Separate | 84.5% | 17.4% | 19.7% | 2.5% | 63.1% | 49.9% | 28.4% | 2.5% | 60.3% |
| | | Borrow | 92.6% | 32.3% | 36.5% | 12.7% | 78.8% | 68.7% | 48.9% | 12.7% | 74.2% |
| Partial alternative 3: only latent class with the ***smallest*** sample size (2) is non-null | | | | | | | | | | | |
| | 3000 | Separate | 2.5% | 17.5% | 2.2% | 2.2% | 2.5% | 2.6% | 2.3% | 13.4% | 17.5% |
| | | Borrow | 1.6% | 0.7% | 0.2% | 1.7% | 1.0% | 0.6% | 0.2% | 4.2% | 0.7% |
| Partial alternative 4: only latent class with the ***largest*** sample size (4) is non-null | | | | | | | | | | | |
| | 3000 | Separate | 2.6% | 2.4% | 2.3% | 87.8% | 2.7% | 2.7% | 2.6% | 14.6% | 87.8% |
| | | Borrow | 3.3% | 0.8% | 0.8% | 71.9% | 2.9% | 2.2% | 1.5% | 9.2% | 71.9% |
| Partial alternative 5: three classes with the ***largest*** sample size (5,1,4) are null | | | | | | | | | | | |
| | 3000 | Separate | 2.7% | 17.4% | 19.8% | 2.4% | 2.2% | 50.7% | 28.8% | 7.1% | 32.9% |
| | | Borrow | 4.8% | 7.6% | 9.9% | 4.8% | 4.6% | 37.6% | 18.0% | 11.9% | 21.6% |
| Partial alternative 6: three classes with the ***largest*** sample size (5,1,4) are non-null | | | | | | | | | | | |
| | 3000 | Separate | 85.0% | 2.4% | 2.2% | 86.3% | 61.7% | 2.2% | 2.2% | 8.8% | 78.1% |
| | | Borrow | 91.3% | 7.5% | 7.7% | 91.7% | 73.6% | 10.3% | 10.0% | 23.1% | 85.9% |

medRxiv

### Supplementary materials – Figures

#### Figure S1 – Conceptual framework of the study design and research stages 1-3

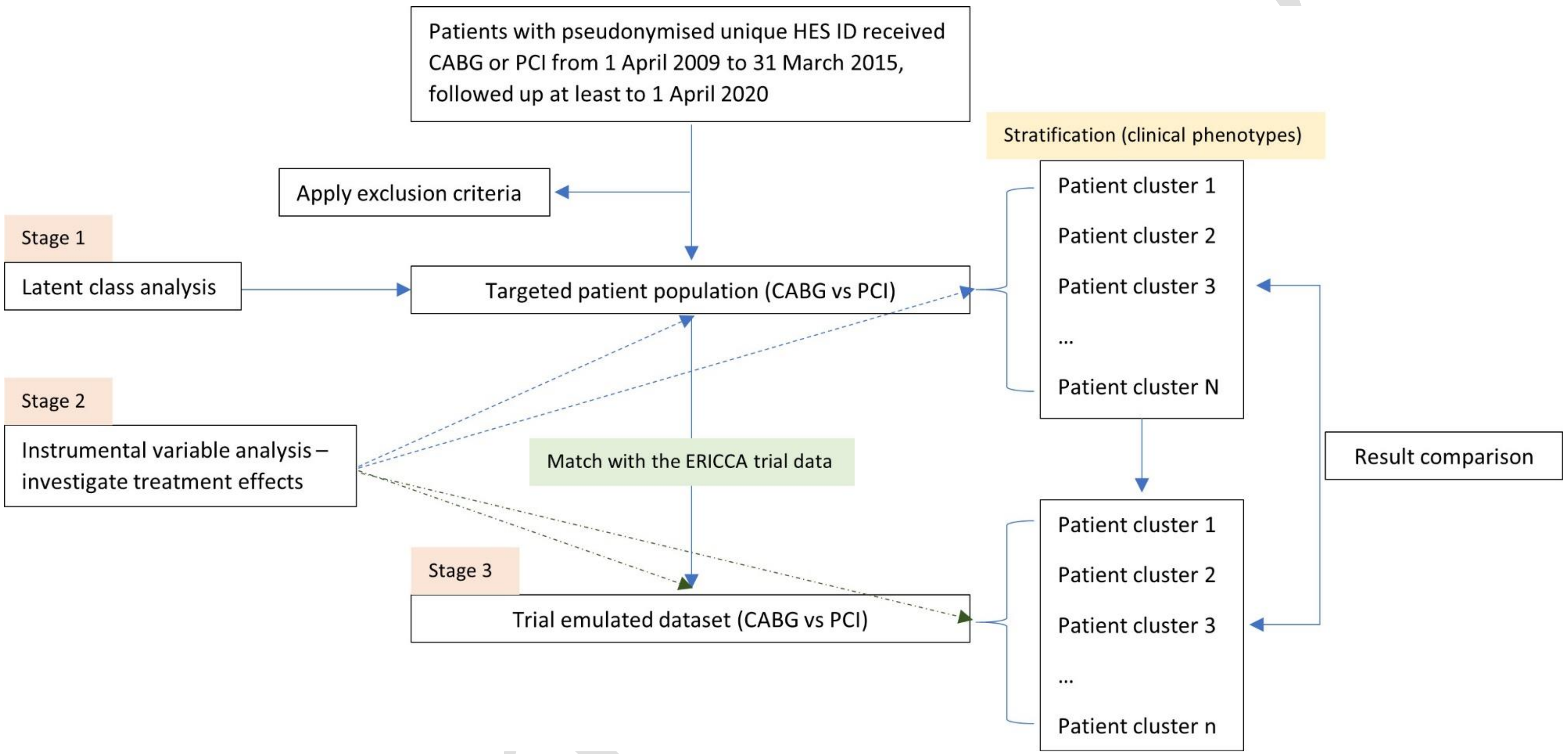

Note: this figure was published in the study design paper in the American Heart Journal (2026, doi: 10.1016/j.ahj.2026.107368).

Figure S2 – Comparing model statistics from two to nine classes using latent class analysis (stage 1)

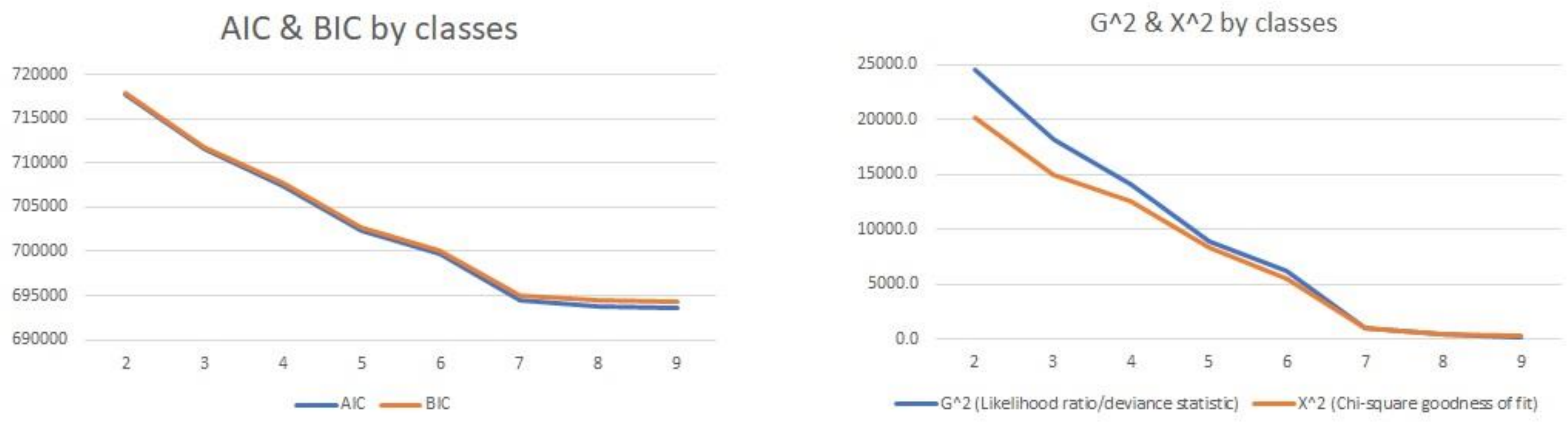

Note: AIC (Akaike Information Criterion), BIC (Bayesian Information Criterion), $G^2$ (Likelihood ratio/deviance statistic), $\chi^2$ (Chi-square goodness of fit)

### Figure S3 – Regional variation in standardised CABG rates in the whole study population (HES) and each latent class (stage 2)

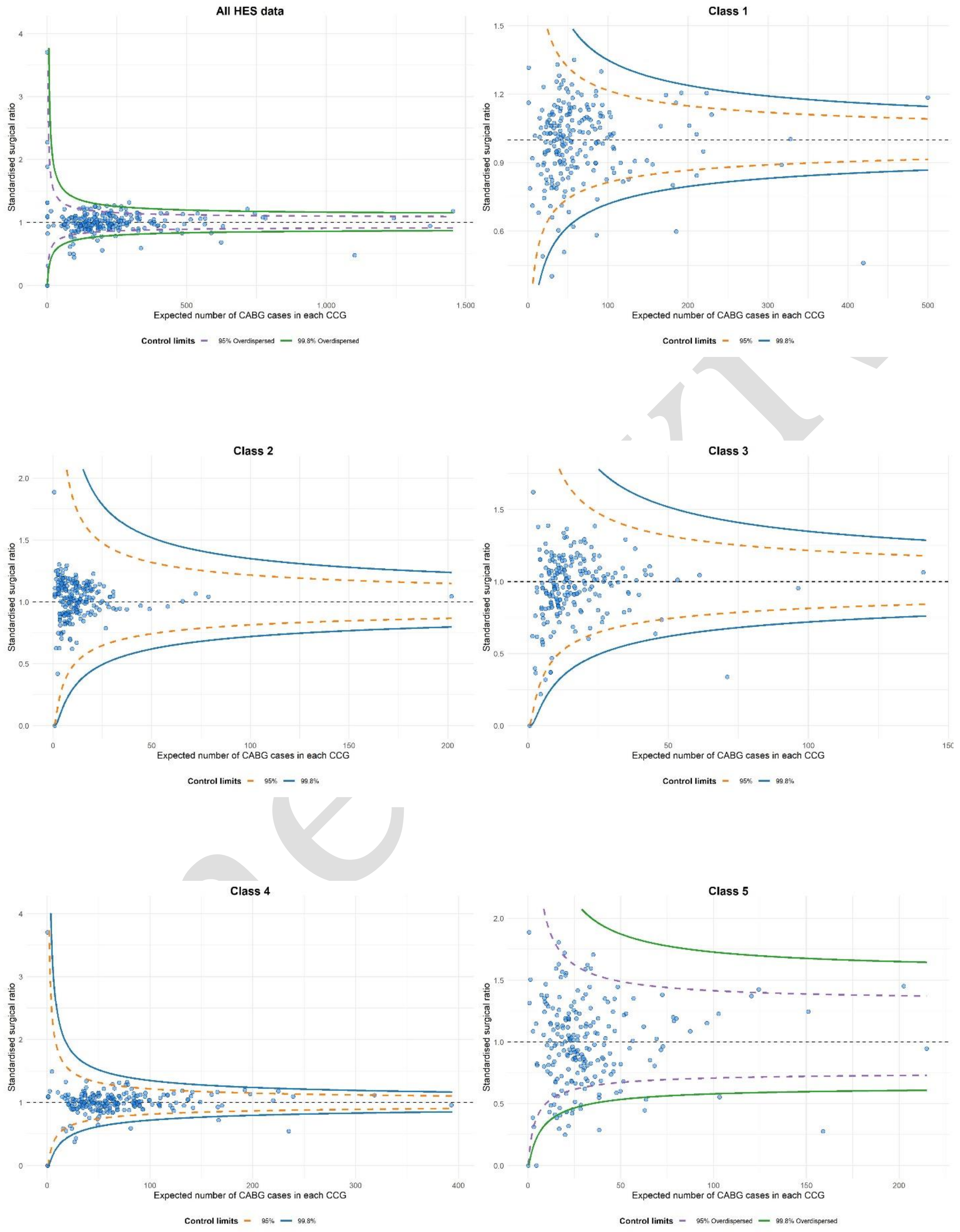

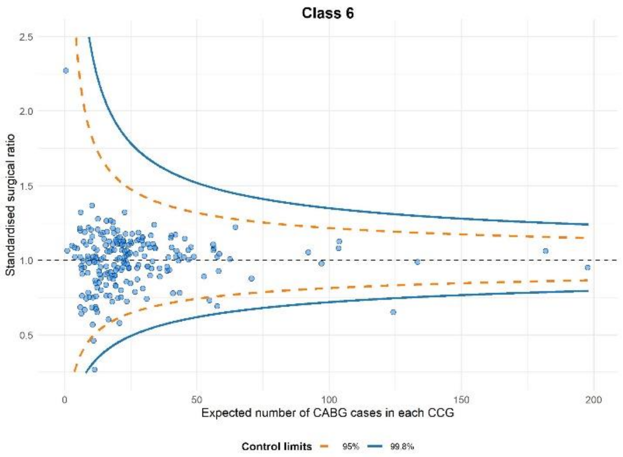
Class 6
Standardised surgical ratio
2.5
2.0
1.5
1.0
0.5
0
50
100
150
200
Expected number of CABG cases in each CCG
Control limits
95%
99.8%

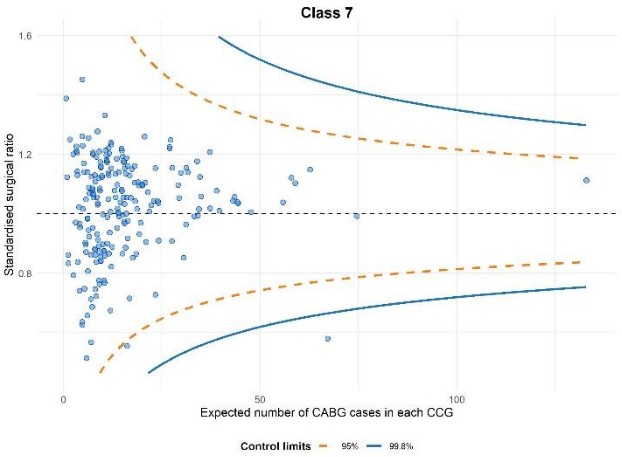
Class 7
Standardised surgical ratio
1.6
1.2
0.8
0
50
100
Expected number of CABG cases in each CCG
Control limits
95%
99.8%

medRxiv

### Figure S4 – Balance of variables before and after propensity score matching between the ERICCA trial and the HES for the emulated trial (stage 3)

Individual participant data from the ERICCA trial 1:1 matched to the HES **CABG** cohort

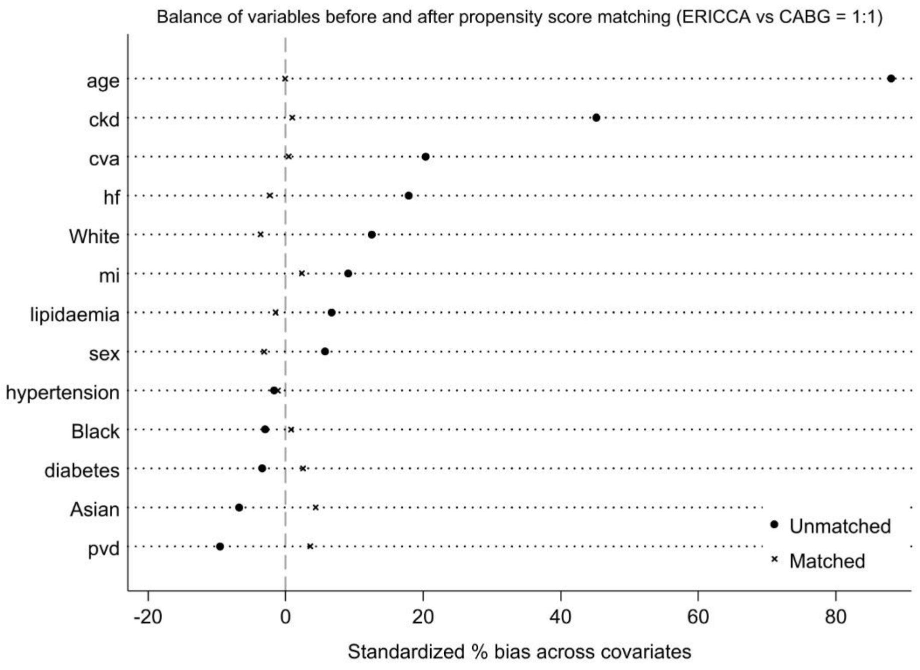

Individual participant data from the ERICCA trial 1:1 matched to the HES **PCI** cohort

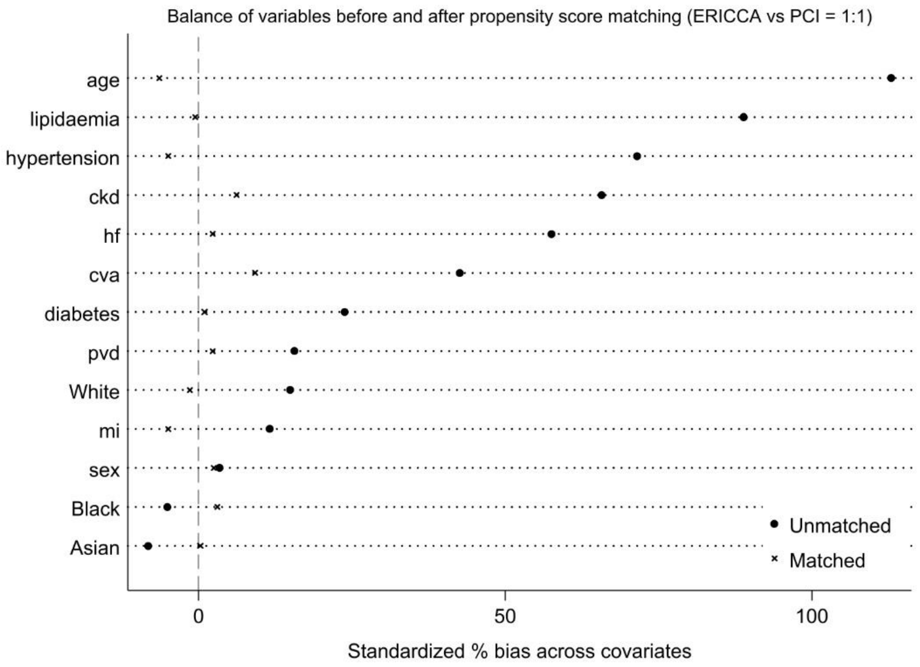

Note: CABG (coronary artery bypass surgery), PCI (percutaneous coronary intervention), HES (hospital episode statistics), ERICCA (doi: 10.1056/NEJMoa1413534)

Figure S5 – Power using Bayesian borrowing or individual classes as separate trials for the target pragmatic trial

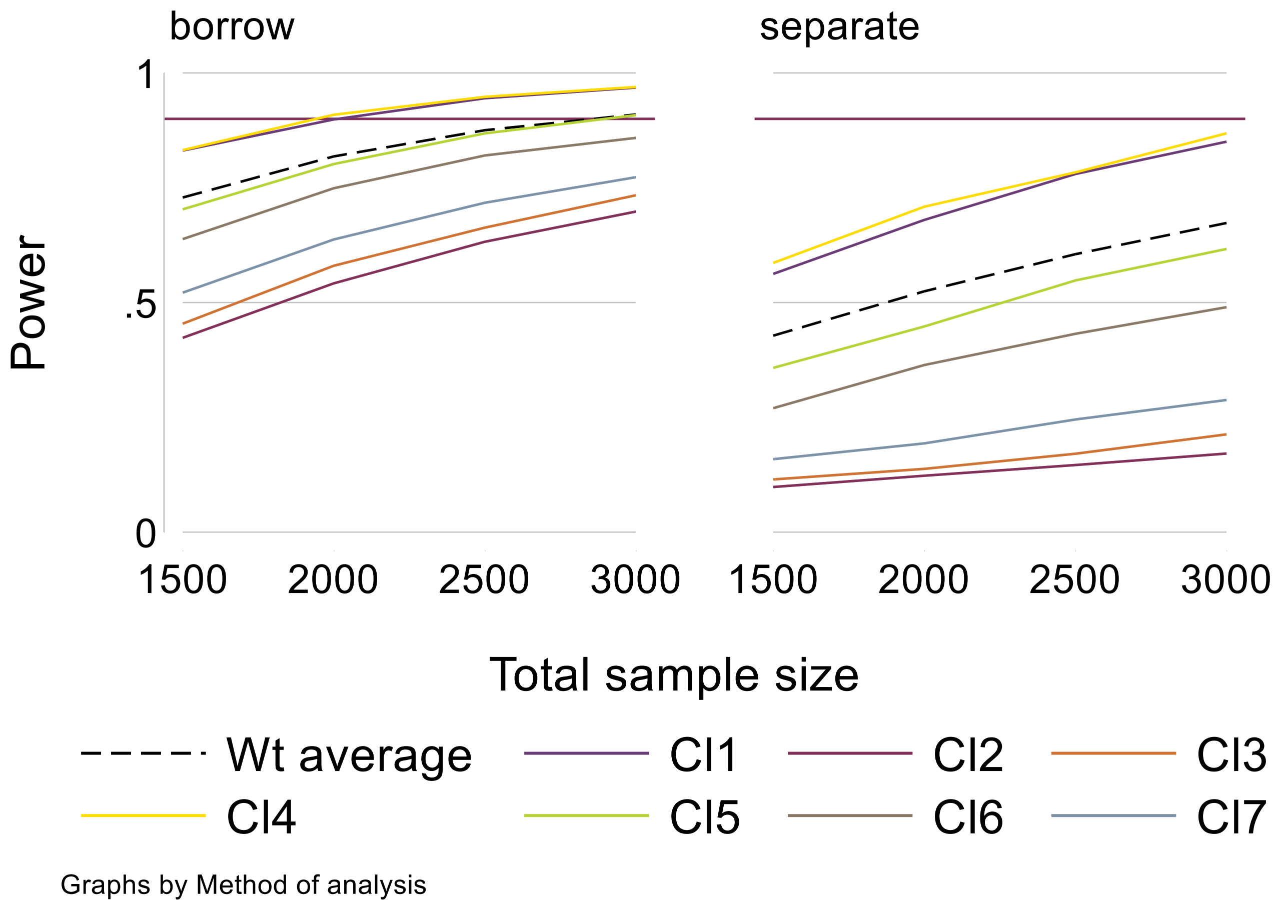

Note: brown horizontal line for 90% power reference

med
